# Preeclampsia Predictor with Machine Learning: A Comprehensive and Bias-Free Machine Learning Pipeline

**DOI:** 10.1101/2022.06.08.22276107

**Authors:** Yun C. Lin, Daniel Mallia, Andrea O. Clark-Sevilla, Adam Catto, Alisa Leshchenko, David M. Haas, Ronald Wapner, Itsik Pe’er, Anita Raja, Ansaf Salleb-Aouissi

## Abstract

Preeclampsia is a type of hypertension that develops during pregnancy. It is one of the leading causes for maternal morbidity with consequences during and after pregnancy. Because of its diverse clinical presentation, preeclampsia is a uniquely challenging adverse pregnancy outcome to predict and manage. In this paper, we explore preeclampsia in a nulliparous study cohort with machine learning techniques to build a model that distinguishes between participants most at risk for morbidity, those with preeclampsia with severe features or eclampsia, and the class of no pregnancy-related hypertension. We curated the dataset for this secondary analysis using only training examples that have all known biomarkers, factors, and placental analytes. We built classification models at discrete time points in pregnancy that combine risk factors for preeclampsia with severe features or eclampsia to help screen cases early in pregnancy. The time points are at 6^0^ − 13^6^ (V1), 16^0^ − 21^6^ (V2), 22^0^ − 29^6^ (V3) weeks gestation and at delivery (V4). We then analyzed the model prediction results and provided an interpretable report of cut-off points of the top contributing risk factors and their impact on prediction. Finally, we identified race-based biases in our models and describe how we mitigate those biases. We evaluated the results of four machine learning algorithms and found that ensemble methods outperformed non-ensemble methods. Random Forest models achieved an area under receiver operating characteristic curve at V1 of 0.68 ± 0.05, V2 of 0.73 ± 0.05, V3 of 0.76 ± 0.04 and V4 of 0.83 ± 0.03. Analyzing the Random Forest models, the features found to be most informative across all visits fall into several broad categories: weight, blood pressure measurements, uterine artery doppler measurements, diet intake and serum biomarkers. We found that our models are biased toward non-Hispanic black participants with a high predictive equality ratio of 1.31. We corrected this bias and reduced this ratio to 1.14. We also evaluated results for predictions of early cases versus late preeclampsia with severe features or eclampsia and found that placental analytes as the top contributors in model feature importance. Random Forest for this analysis achieved an area under receiver operating characteristic curve at V1 of 0.63 ± 0.11, V2 of 0.79 ± 0.11, V3 of 0.83 ± 0.08 and V4 of 0.84 ± 0.09. Our experiments suggest that it is important and possible to create screening models to predict the participants at risk of developing preeclampsia with severe features and eclampsia. The top features stress the importance of using several tests, in particular tests for biomarkers and ultrasound measurements. The models could be used as a screening tool as early as 6-13 weeks gestation to help clinicians identify participants who may subsequently develop preeclampsia, confirming the cases they suspect or identifying unsuspected cases. The proposed approach is easily adaptable to address any adverse pregnancy outcome with fairness.

## Introduction

Preeclampsia (PE) is one of the leading causes for maternal morbidity, with consequences during and after pregnancy. ^1^ Ensuring optimal patient outcomes requires robust prediction models for PE risk, with a particular emphasis on early detection. However, PE poses significant diagnostic and prognostic difficulties given its variable presentations, in terms of clinical indications, speed of development and timing, as well as its unknown causes. PE might evolve slowly and remain mild or might quickly present severe complications leading to what is known as PE with severe features (sPE). ^2^ Moreover, there are two sub-categories: early onset PE requiring delivery before 34 weeks, and late onset thereafter. While early onset of PE is associated with a higher incidence of adverse pregnancy outcomes, understanding the relationship between the early and late onset of PE has proven to be challenging. ^3 4^ Some researchers treat them as distinct, but work by Poon et al ^3^ treats the condition as a spectrum, best represented by a survival time model. Beyond this, the presence of seizures that cannot be attributed to any other underlying condition in a patient diagnosed with PE would be categorized as Eclampsia (E). ^2^

Though a full understanding of PE remains elusive, there exists a rich literature on risk factors for, and indicators of, PE. Biochemical and biophysical markers can have an added benefit for screening for PE when used in combination with clinical characteristics taken from medical history, demographics, clinical measurements, etc. ^3 5 6 7 8^ Research ^3 9 10 11^ has suggested placental growth factor (PlGF), soluble Flt-1 (sFlt-1), pregnancy-associated plasma protein A (PAPP-A) and ultrasound measurements as clinical factors that are significant in signaling an increase in the risk of PE.

Applying this significant volume of knowledge to prediction is pertinent. The objective of this study was to build bias-free machine learning classifiers at various discrete points in pregnancy that combine most previous known risk factors for and indicators of sPE and E, and can help screen for cases early in pregnancy.

## Materials and Methods

### Study population

The retrospective study cohort to which we applied machine learning is the Nulliparous Pregnancy Outcomes Study: Monitoring Mothers-to-be (nuMoM2b) study, ^12^ which contains information from eight clinical sites across the US. Participants gave written informed consent and institutional review board approval was obtained at all sites. The study contains a wide array of information collected for nulliparous participants across four visits, three corresponding roughly to the three trimesters (V1-V3) and a delivery visit (V4). At V1 and V2, maternal serum was collected, enabling a limited follow-up nuMoM2b sub-study, conducted to understand the relationship between placental analytes and a set of adverse pregnancy outcomes (APOs). Figure 1 describes in detail the number and categories of features selected.

**Figure 1:**
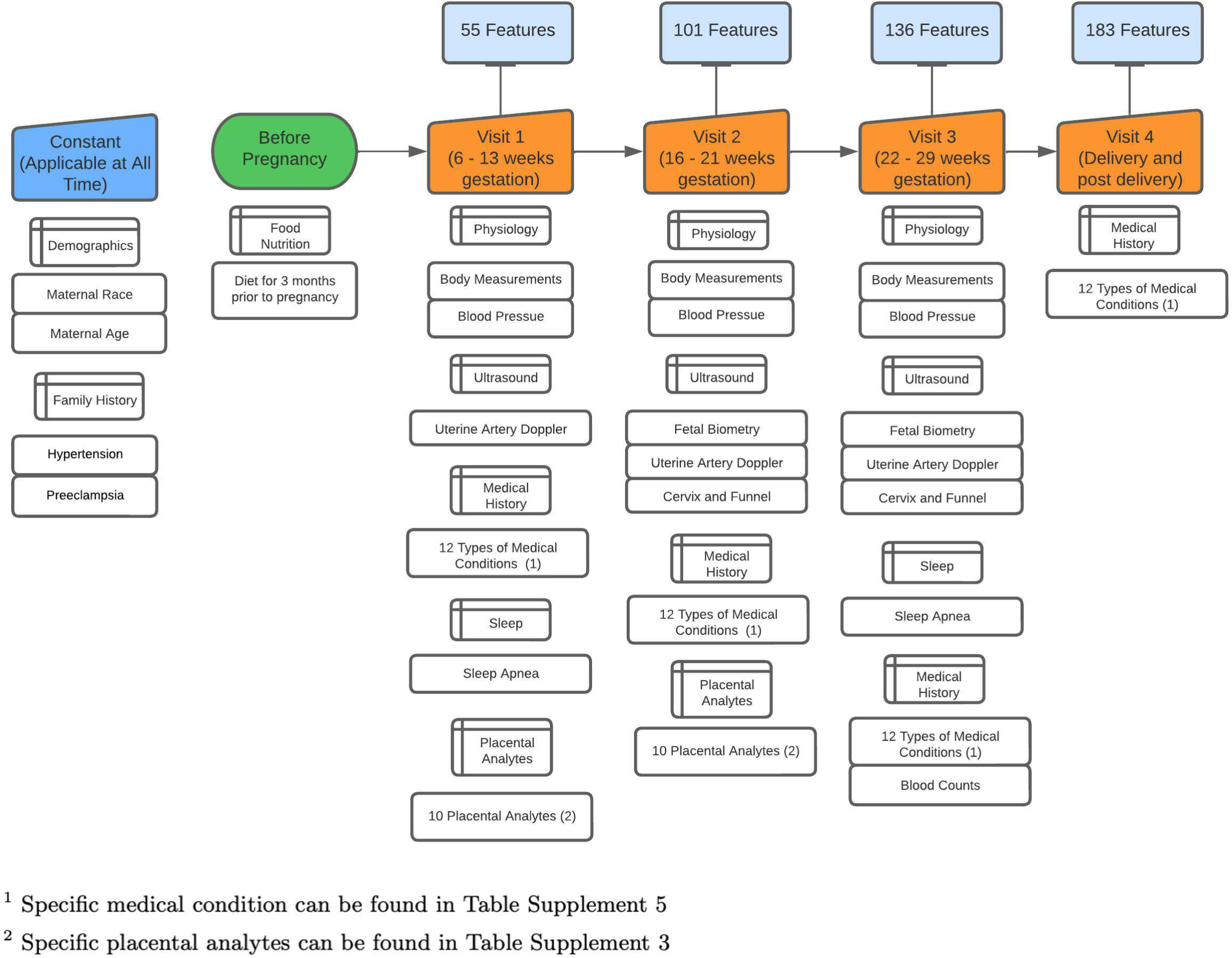
Data process timeline This figure shows the gestational weeks at each visit. For each visit, the number of features at that visit is listed and the category of new feature included is also shown.

To focus on those most at risk, participants with chronic hypertension condition were removed, as were those with mild preeclampsia, superimposed preeclampsia and new onset hypertension. There are no cases of fetal demise at <20 weeks cases in the final study cohort. We preserved 36 cases of stillbirth, but these all fell into the no pregnancy related hypertension (NPH) category.

### Study outcome

The labeling of sPE employed was that of the nuMoM2b study. Appendix (Supplement Figure 2) contains a flowchart indicating the study diagnostic criteria for sPE. The nuMoM2b dataset also contained labels in accordance with the ACOG criteria published in 2013. However, because this was an adaptation somewhat limited by the study data collection practices and initial testing of the proposed pipeline with this labeling indicated results very similar to that achieved with the nuMoM2b criteria, the nuMoM2b labeling was selected as the target for this study.

### Study population characteristics

Statistics were gathered for all features across visits spanning multiple categories. We compared sPE + E versus NPH and early sPE versus late sPE + E patients. For continuous factors, Kolmogorov-Smirnov test was used to test for normality. If the factor is normally distributed, we used Welch’s t-test, otherwise we used the Mann–Whitney U test. For categorical features, we used the Chi Square test or Fisher Exact test (2 categories).

### PEPrML pipeline

Our **P**re**E**clampsia **Pr**edictor with **M**achine **L**earning (PEPrML) pipeline produces machine learning capable models that are explainable and trustworthy. Classifiers to predict sPE+E versus NPH and early sPE versus late sPE+E were learned for every visit. Categorical features were one-hot encoded. For continuous variables with missingness, mean imputation was used. We used a cross-validation strategy that uses 60-20-20 percent train, validation, and test splits, conducting 100 trials for each of the experiments. We balanced the ratio of control versus targets by undersampling in the training and test sets, as this introduces less overfitting, leads to a faster training time and avoids an over-inflated Area Under the ROC curve (AUC). We experimented with logistic regression (LR), support vector machines (SVM), random forest (RF) and eXtreme Gradient Boosting (XGBoost). ^13^ For RF and XGBoost we extracted the interpretable feature importance rankings, identifying the top factors for which to generate partial dependence plots (PDPs). ^14^ PDPs display the marginal effect that one or two features have on the predicted outcome of a given model, allowing us to advance our understanding of the outcome. For the RF model we calculated the Equal Opportunity Ratio (EOR), ^15^ Predictive Parity Ratio (PPR), ^16^ Predictive Equality Ratio (PER), ^16^ Accuracy Equality Ratio (AER), ^17^ and Statistical Parity Ratio (SPR), ^16^ and mitigate the race-based biases using *Ceteris Paribus* Cutoff Plot. For details please refer to the Appendix.

## Results

### Study population characteristics

1,758 participants were selected as the final study cohort. Among these, 5 developed E, 273 developed sPE, of which 58 (∼21%) were early onset (<34 weeks), and 215 (∼79%) were late onset. The remaining 1,480 patients were NPH (Supplement Figure 1).

Among participants with sPE+E and NPH, features that are highly significant (P<0.001) are BMI, blood pressure measurements (V1-V3), patient weights (V1-V3), waist and neck circumference, and uterine artery doppler measurements (V1, V2). Six analytes: endoglin (V2), inhibin A (V2), ADAM12 (V2), VEGF (V1), PlGF (V1, V2), and sFLT1-to-PlGF ratio (V2), are also highly significant. Only two medical conditions appear highly significant for comparing participants with sPE and NPH: diabetes (V1, V2), hypertension (V3, V4).

Considering early sPE versus late sPE+E, diastolic blood pressure (V3), mean arterial pressure (MAP) (V3) and for most of the uterine artery doppler measurements, V2 and V3 measurements are highly significant. Five analytes: inhibin A (V2), ADAM12 (V2), PAPP-A (V1), PlGF (V1, V2), and sFLT1-to-PlGF ratio (V2), are also highly significant. Only one medical condition, kidney disease (V1, V2, V4) is highly significant.

For a detailed summary please refer to the Appendix (Supplementary Tables 1 - 5).

### Model performance

A summary of performance results for sPE+E versus NPH can be found in Figure 2. Results in Figure 2.a indicate that prediction capabilities increase with gestational age. RF models achieved an AUC of V1 of 0.68 ± 0.05, V2 of 0.73 ± 0.05, V3 of 0.76 ± 0.04 and V4 of 0.83 ± 0.03. Detailed performance metrics can be found in Table 1. Further breakdown of the performance is offered in Tables Supplement Table 6 and Supplement Table 7, which summarize results for predicting early and late onset, versus NPH, respectively.

**Figure 2:**
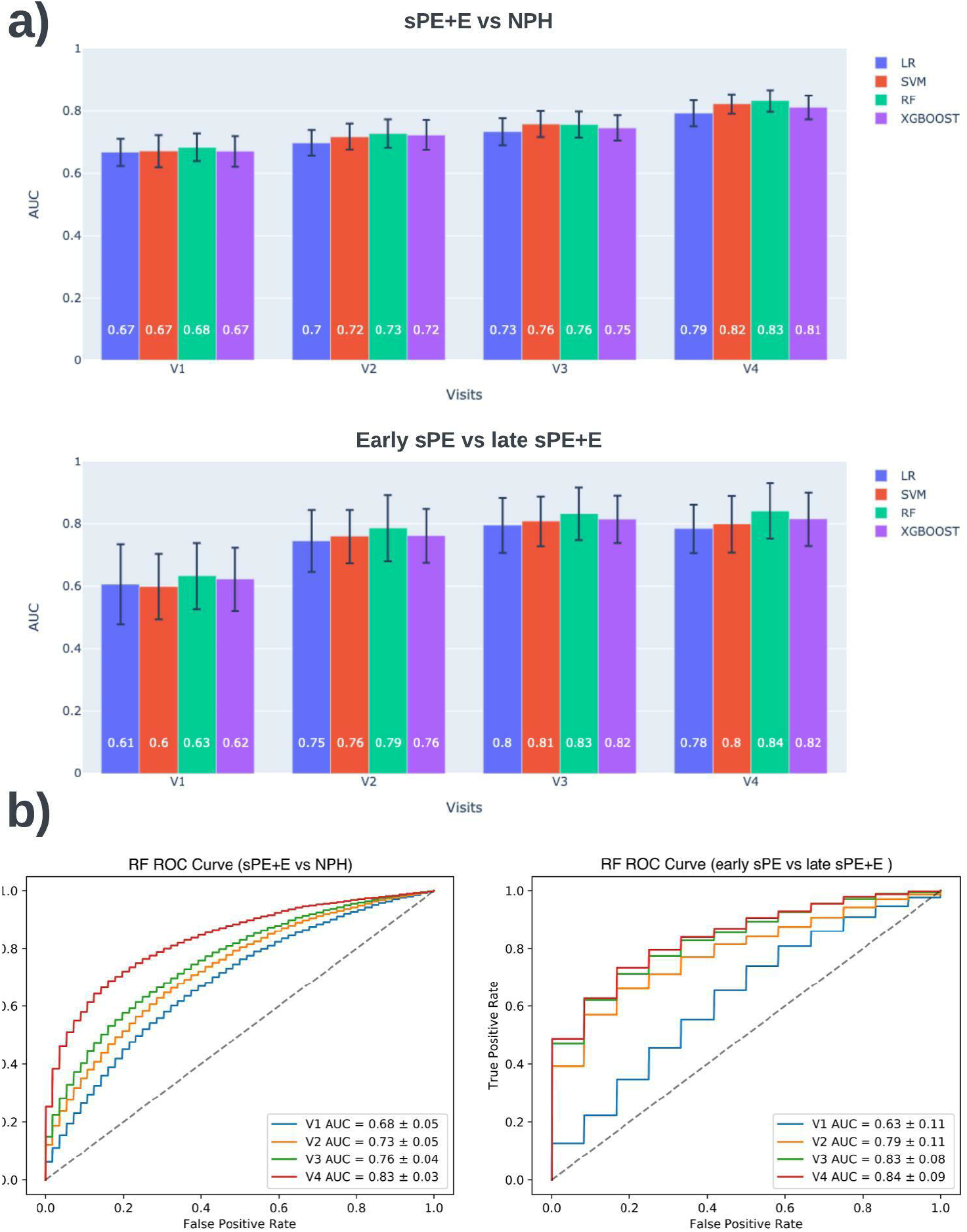
sPE+E vs NPH and early sPE vs late sPE+E model performance. a) Average AUC for 100 trails per visit for 4 classifiers. b) RF classifier has best performance across visits for both comparisons. The ROC curve demonstrated the tradeoff between the true positive rate versus false positive rate. This summarizes the results for 100 trails.

**Table 1:**
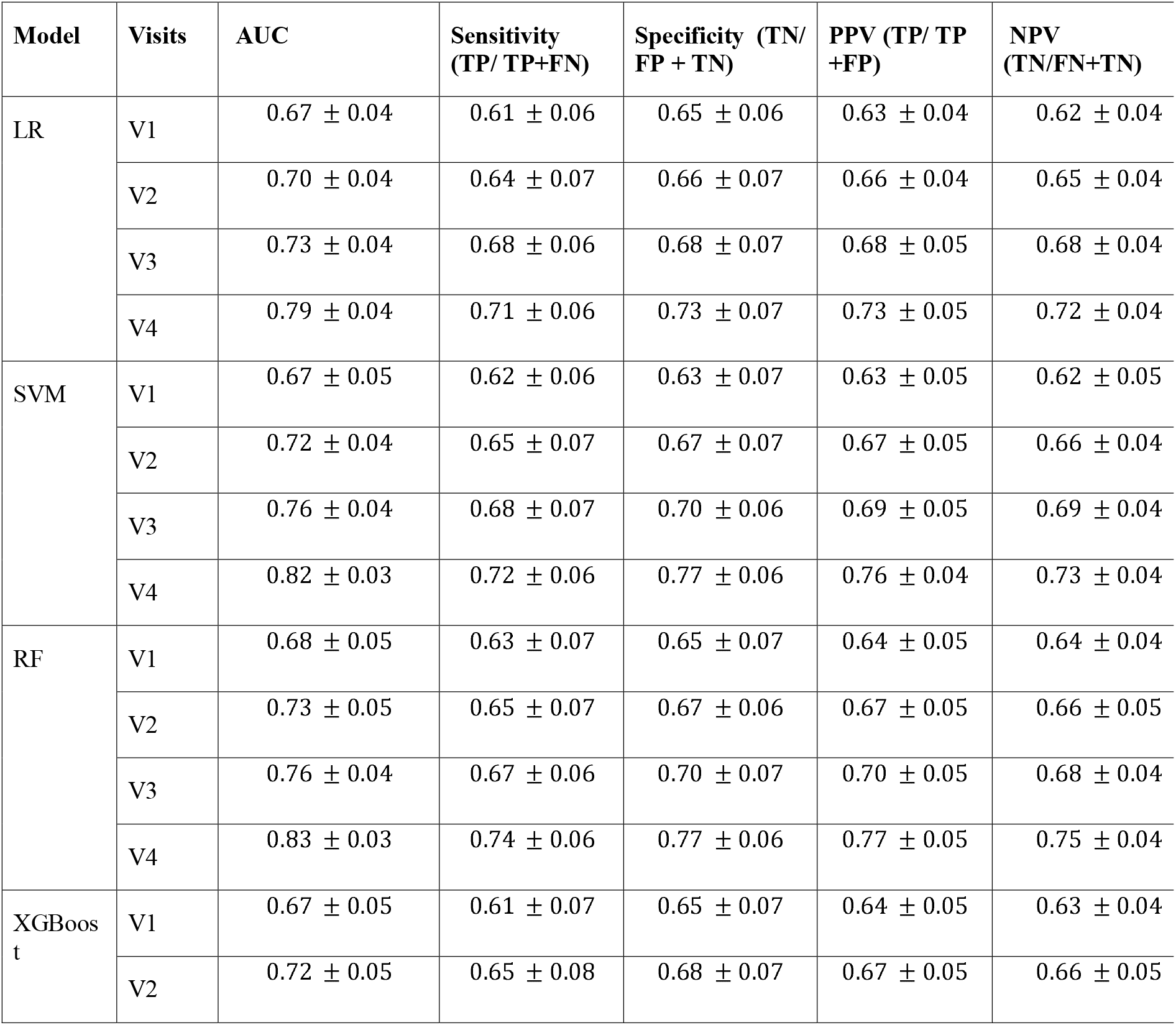
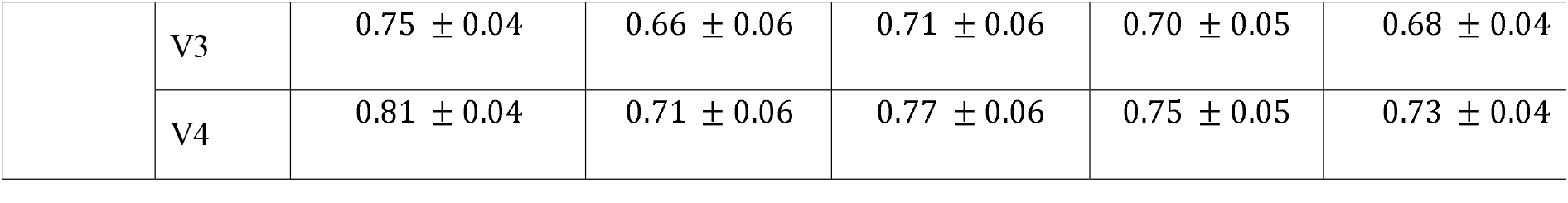
Detailed summary of sPE+E vs NPH model performance per visit for 4 classifiers.

Prediction power of the model for early onset preeclampsia is higher than for late onset as demonstrated by the two tables. Across the board all metrics have higher value, but variance is also higher for these values, which is mostly likely due to the smaller set of cases with early onset sPE. We learned classifiers to directly predict early sPE vs late sPE+E to better understand what enabled this performance. A summary of performance results for early sPE vs late sPE+E can be found in Figure 2. Again, performance increased with gestational age and RF models performed the best, obtaining an AUC of 0.63 ± 0.11 for V1, 0.79 ± 0.11 for V2, 0.83 ± 0.08 for V3 and 0.84 ± 0.09 for V4. Detailed performance metrics can be found in Supplement Table 8.

### Interpreting sPE+E vs NPH model

The feature importance lists for V1, V2 and V3, where the prediction task is prognosis, are given in Supplement Figure 3, Figure 3.a, and Supplement 4, respectively, enabling better understanding of the key features that contribute to the RF and XGBoost decision processes. For V1, the top 5 features are BMI, SBP, MAP, waist circumference and endoglin. For V2, the top 5 features are BMI, PlGF (V2), SBP (V2, V1), MAP (V2).

**Figure 3:**
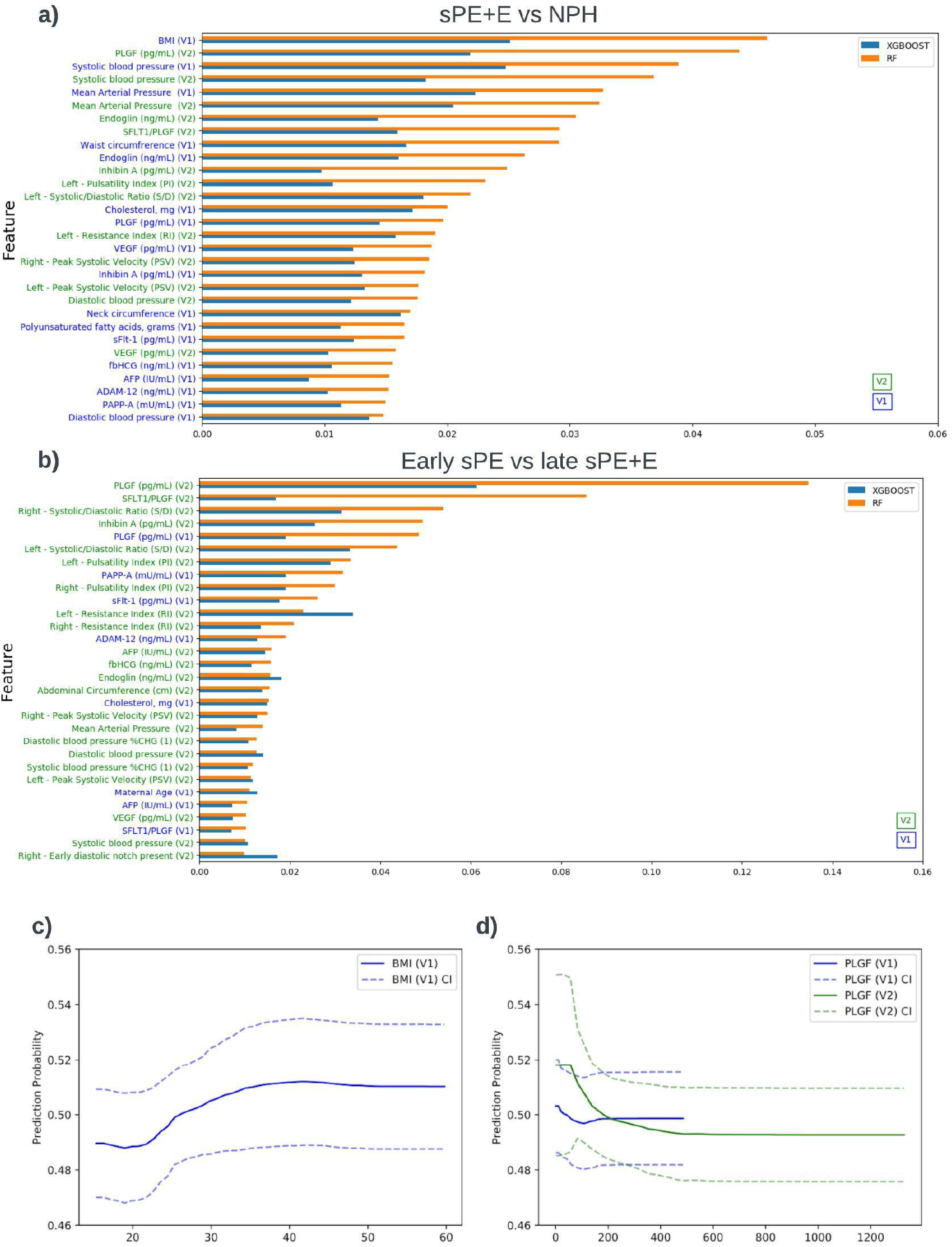
Interpreting machine learning model for sPE+E vs NPH and early sPE vs late sPE+E a) V2 features importance for sPE+E vs NPH model, b) V2 feature importance for early sPE vs late sPE+E, c - d) PDP for BMI and PlGF based on model build for sPE+E vs NPH

The PDP for BMI shown in Figure 3.c indicates a significant risk increase in sPE+E at around 22.41 *km*/*m*^2^. We see a significant increase in the risk of sPE+E with a systolic reading of 110 mmHg or higher, and by Visit 2 this number drops to 102 mmHg. (Supplement Figure 5.a) The diastolic reading did not exhibit such a pronounced increase in the risk of sPE+E, but we did observe a slight increase above 78 mmHg. Looking at the MAP at Visit 1, Supplement Figure 5.b, we see an increase in risk at 82.67 mmHg. There is a sharp increase in the predictive risk for sPE+E observed in the PDP for PlGF at Visit 1 for measurements less than 100 pg/mL.

### Racial Fairness in sPE+E vs NPH model

We observed that our model for predicting sPE+E vs NPH is biased mainly against Black participants. Using the White race as the reference race, we identified that the predictive equality ratio for Black participants (1.31) is high according to the four-fifths rule.

To address this problem, we plotted the *ceteris paribus* Cutoff plot of the parity loss for the Black sub-population to determine the optimal confidence threshold for prediction. Adjusting the threshold accordingly mitigated the over-prediction of PE occurrence by our model for Black participants, reducing the predictive equality ratio for Black participants from 1.31 to 1.14 (Figure 4).

**Figure 4:**
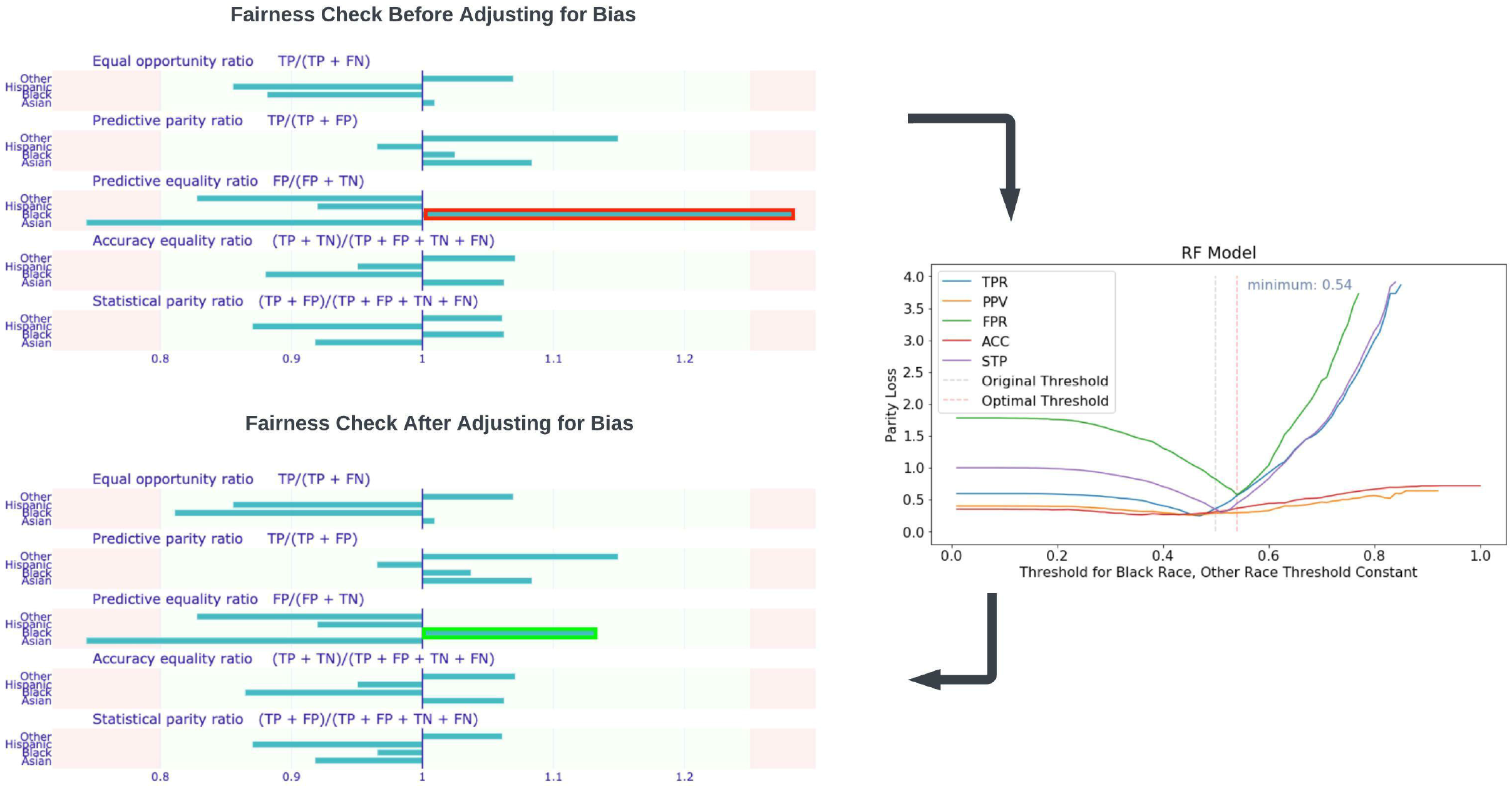
Fairness check for sPE+E vs NPH mode The threshold set based on the four-fifth rule are 0.8 and 1.25. *Ceribus Paribus* plot was used to adjust prediction threshold for the Black population.

## Comment

### Principal Findings

The results presented here demonstrate that it is possible to automatically learn RF models with superior, well-rounded performance for early prediction of preeclampsia at multiple time points throughout pregnancy, with minimal preprocessing of data, feature engineering or feature selection. Exhibiting no steep PPV - Sensitivity trade-off without any tuning, RF increases in performance by all metrics at each new visit, as more information becomes available. The feature importance plots confirm existing knowledge about known predictive features such as blood pressure, uterine artery blood flow, and placental analytes, and identifies features not commonly referenced in the prediction literature, such as Endoglin, Cholesterol, and Inhibin A. Review of RF fairness metrics indicated a correctable bias against Black participants.

### Results in the Context of What is Known

Our study confirmed that blood pressure and placental analytes were significant in predicting PE across study visits. ^18 19 20^ Results of our statistical tests deviate from other works ^3 11 21^ in that risk factors such as maternal age, race, sleep apnea and history of PE were not significant. Thus, care must be taken in comparing the model performance presented here for the nuMoM2b dataset with other studies given that the nuMoM2b dataset characterizes demographically diverse nulliparous mothers with unknown risk for PE at the time of first prediction while the target label is strictly focused on sPE+E criteria.

JHee et al ^22^ and Maric et al ^23^ are two studies with close relevance to our work although both use regional datasets. JHee et al focused only on the prediction of late onset PE. They found that a stochastic gradient boosting model yielded the highest performance (AUC 0.924). Maric et al ^23^ presented models to predict all PE cases and early onset PE, finding that GBTree yielded the highest performance for the prediction of all PE cases (AUC 0.79) and for the prediction of early onset PE (AUC 0.89). However, their test set may have been imbalanced, given the low PPV and high AUC. Our findings that (a) prediction of early onset sPE vs NPH generated a higher AUC than late sPE vs NPH and (b) ensemble methods, viz. RF and XGBoost ^24^, are the top performers is consistent with the literature. ^22 23 25^ This may be due to the ensemble nature, and the ability of the underlying model, decision trees, to capture some of the subtle distinctions between the varied and poorly understood subgroups of preeclampsia patients. ^26^ Unlike the above works, our study did not include chronic hypertensive patients and the presence of chronic hypertension was not included as a feature since it serves as a proxy for superimposed PE.

The PDP for BMI, a well-known risk factor for PE, shown in Figure 4.a indicates a significant risk increase in PE around 22.41 km/m^2^. While this cutoff point is still within the healthy range (18.5-24.9 kg/m^2^), ^27^ this measure might be factored into future ACOG guidelines. Furthermore, PDPs for various placental analytes indicate that a decreased level of PlGF during the first and second trimesters precede the onset of PE. ^3 28 29^ Agrawal et al ^30^ found that the predictive values were highest for PlGF levels between 80 and 120 pg/mL, which coincides with the sharp increase in the predictive risk for PE observed in the PDP for PlGF at Visit 1 for measurements less than 100 pg/mL. MacDonald et al ^31^ suggested a sFlt-1:PlGF ratio > 33.4 which is in agreement with our PDP in Supplement Figure 7. Levine at al ^32^ found that levels of endoglin at 25 through 28 weeks of gestation were found to be significantly higher (8.5 ng/mL) in term PE patients. We observe this same cut-off value in the PDP in Figure 4.e which shows a pronounced increase in the risk of PE at around 9 ng/mL at V1, albeit occurring much earlier, at 6-13 weeks of gestation. That analytes such as PlGF, unlike blood pressure, were consistently important across the original model and the early vs late model (Figure 4), indicates their predictive power, particularly that of a *rule out* ability for early onset. ^5 25^

### Clinical Implications

This study demonstrates the utility of early and multiple time point screening for PE. It shows early measurement of blood pressure can serve as proxy for risk of high blood pressure later in pregnancy. Also, information about placental analytes, which can potentially be gathered at a reasonable cost tradeoff between assessment and hospitalization, ^5^ allow predictions that strongly surpass the accuracy of an ACOG recommended medical history review. ^33^ Further validation is required for the proposed separate models for multiple time points to ensure consistency of prediction: a patient identified as high risk early in pregnancy should not be deemed low risk later without sufficient explanation.

Fairness metrics and analysis of causes for biases should become part of standard practice in model validation. We hypothesize that the bias against the Black participants may have been caused by the limited sample size, which is skewed disproportionately towards White mothers, and the potentially inappropriate higher representation of Black population among the sPE+E class than the NPH class (20.9% vs. 13.8% respectively). While the cost of a false negative diagnosis for maternal and fetal health is very high, the stress, costs, and possibly inappropriate treatment of a false positive should not be ignored.

### Research Implications

Distinguishing between sPE+E and NPH is a critical task, but the binary labels pose a challenge. The latter group undoubtedly contains different subgroups and phenotypes of preeclampsia and learning to make these distinctions will have the dual benefit of enhancing our understanding of preeclampsia and allowing for better predictive performance. Thus, moving beyond the initial literature-inspired feature set to a broader set of features will be the target of future work. Furthermore, temporal features capturing change between clinical measurements at different visits, will be investigated, as this may enhance the quality of prediction at the second and third time points. ^26^ This would enable more timely monitoring and treatment of late onset preeclampsia.

A more significant departure will involve re-framing the prediction task. Compelling arguments have been made that preeclampsia is best interpreted as a syndrome rather than a disease. ^34 26^ Label difficulties have led at least one study of short term preeclampsia screening to focus on a label that consists of the presence, or not, of at least one of multiple maternal or fetal adverse outcomes. ^25^ Considering whether this or a continuous spectrum is more suitable, will be the subject of future work.

### Strengths and Limitations

For this initial study, a reasonable set of features identified in the related medical literature was employed, but this can be expanded without issue. The decision not to use other transformations such as multiple of the median (MoM) values, means that the proposed pipeline is dataset and population agnostic, and can easily be adapted for more data or a different task. Use of the nuMoM2b data represents an exciting opportunity to learn from a sizeable sample of U.S. mothers that is more diverse than other similar studies and that has been captured in a longitudinal study with a considerable number of features. ^4 25 35^ The occurrence rate of PE in this study was consistent with reported rates. ^5 36^ However, this meant that even with such a sizeable sample, the analysis was limited to only a couple of hundred sPE+E cases. The sub-study was also not without its limitations: analytes were only available for V1 and V2, and ultrasound data was only generally available beginning at V2.

One noticeable limitation of the study is the limited cases of existing medical conditions in participants of the placental analytes sub-study. This low presence can cause the model to attribute less importance to these risk factors, while these could be crucial in clinical application. Lastly, our study only focuses on comparing patients with sPE+E and NPH, without addressing those patients that developed superimposed PE, PE with mild features, or only hypertension.

## Conclusions

Our experiments suggest that it is important and possible to create screening models to predict the participants at risk of developing preeclampsia with severe features and eclampsia. The top features stress the importance of using several tests, in particular tests for biomarkers and ultrasound measurements. The models could be used as a screening tool as early as 6-13 weeks gestation to help clinicians identify participants who may subsequently develop preeclampsia, confirming the cases they suspect or identifying unsuspected cases. The proposed approach is easily adaptable to address any adverse pregnancy outcome with fairness.

## Data Availability

All data produced in the present work are contained in the manuscript

## Acknowledgments

The NuMoM2b Study (Nulliparous Pregnancy Outcomes Study: Monitoring Mothers-to-Be) was supported by grant funding from the Eunice Kennedy Shriver National Institute of Child Health and Human Development (NICHD): U10 HD063036; U10 HD063072; U10 HD063047; U10 HD063037; U10 HD063041; U10 HD063020; U10 HD063046; U10 HD063048; and U10 HD063053.

This work also was awarded innovation and health disparities prizes in NICHD’s Decoding Maternal Morbidity Data Challenge.

## Reference

1. Resnik R, Creasy RK, Iams JD, Lockwood CJ, Moore T, Greene MF. Creasy and Resnik’s maternal-Fetal medicine: Principles and practice E-book: Elsevier Health Sciences; 2008.

2. Creasy RK, Resnik R, Iams JD. Maternal Fetal Medicine : Principles and Practice: Fifth edition. Philadelphia : W.B. Saunders Co.; 2004.

3. Poon LC, Nicolaides KH. Early prediction of preeclampsia. Obstetrics and Gynecology International. 2014;2014.

4. Wójtowicz A, Zembala-Szczerba M, Babczyk D, Kołodziejczyk-Pietruszka M, Lewaczyńska O, Huras H. Early-and Late-Onset Preeclampsia: A Comprehensive Cohort Study of Laboratory and Clinical Findings according to the New ISHHP Criteria. International Journal of Hypertension. 2019;2019:1–9.

5. Sroka D, Verlohren S. Short Term Prediction of Preeclampsia, 2021.

6. Facco FL, Lappen J, Lim C, Zee PC, Grobman WA. Preeclampsia and sleep-disordered breathing: A case-control study. Pregnancy Hypertension: An International Journal of Women’s Cardiovascular Health. April 2013;3:133–139.

7. Eskild A, Vatten LJ. Abnormal bleeding associated with preeclampsia: a population study of 315,085 pregnancies. Acta Obstetricia Et Gynecologica Scandinavica. 2009;88:154–158.

8. Conde-Agudelo A, Villar J, Lindheimer M. Maternal infection and risk of preeclampsia: systematic review and metaanalysis. American Journal of Obstetrics and Gynecology. January 2008;198:7–22.

9. Fox R, Kitt J, Leeson P, Aye CYL, Lewandowski AJ. Preeclampsia: risk factors, diagnosis, management, and the cardiovascular impact on the offspring. Journal of clinical medicine. 2019;8:1625.

10. Karumanchi SA, Epstein FH. Placental ischemia and soluble fms-like tyrosine kinase 1: cause or consequence of preeclampsia? Kidney international. 2007;71 10:959–61.

11. Verlohren S, Herraiz I, Lapaire O, et al. New Gestational Phase–Specific Cutoff Values for the Use of the Soluble fms-Like Tyrosine Kinase-1/Placental Growth Factor Ratio as a Diagnostic Test for Preeclampsia. Hypertension. 2014;63:346–352.

12. Haas DM, Parker CB, Wing DA, et al. A description of the methods of the Nulliparous Pregnancy Outcomes Study: monitoring mothers-to-be (nuMoM2b). American journal of obstetrics and gynecology. 2015;212:539–e1.

13. Chen T, Guestrin C. Xgboost: A scalable tree boosting system. Paper presented at: Proceedings of the 22nd acm sigkdd international conference on knowledge discovery and data mining, 2016.

14. Goldstein A, Kapelner A, Bleich J, Pitkin E. Peeking Inside the Black Box: Visualizing Statistical Learning with Plots of Individual Conditional Expectation

15. Hardt M, Price E, Srebro N. Equality of opportunity in supervised learning. Advances in neural information processing systems. 2016;29.

16. Chouldechova A. Fair prediction with disparate impact: A study of bias in recidivism prediction instruments. Big data. 2017;5:153–163.

17. Verma S, Rubin J. Fairness definitions explained. Paper presented at: 2018 ieee/acm international workshop on software fairness (fairware), 2018.

18. Sibai BM, Ewell M, Levine RJ, et al. Risk factors associated with preeclampsia in healthy nulliparous women. American journal of obstetrics and gynecology. 1997;177:1003–1010.

19. Smith GCS, Stenhouse EJ, Crossley JA, Aitken DA, Cameron AD, Connor JM. Early pregnancy levels of pregnancy-associated plasma protein a and the risk of intrauterine growth restriction, premature birth, preeclampsia, and stillbirth. The Journal of clinical endocrinology and metabolism. 2002;87 4:1762–7.

20. McLaughlin K, Snelgrove JW, Audette MC, et al. PlGF (Placental Growth Factor) Testing in Clinical Practice: Evidence From a Canadian Tertiary Maternity Referral Center. Hypertension. 2021;77(6):2057–2065.

21. Phan K, Pamidi S, Gomez YH, et al. Sleep-disordered breathing in high-risk pregnancies is associated with elevated arterial stiffness and increased risk for preeclampsia. American Journal of Obstetrics and Gynecology. December 2021.

22. Jhee JH, Lee S, Park Y, et al. Prediction model development of late-onset preeclampsia using machine learning-based methods. PLoS ONE. 2019;14.

23. Marić I, Tsur A, Aghaeepour N, et al. Early prediction of preeclampsia via machine learning. American Journal of Obstetrics & Gynecology MFM. 2020;2:100100.

24. Couronné R, Probst P, Boulesteix AL. Random forest versus logistic regression: a large-scale benchmark experiment. BMC bioinformatics. 2018;19:1–14.

25. Schmidt MLJ, Rieger MO, Neznansky MM, et al. A machine-learning based algorithm improves prediction of preeclampsia-associated adverse outcomes. American Journal of Obstetrics and Gynecology. 2022.

26. Myatt L. The prediction of preeclampsia: the way forward. Am J Obstet Gynecol. 2022;226:S1102–S1107.e8.

27. Rasmussen KM, Yaktine AL. Weight Gain During Pregnancy: Reexamining the Guidelines, 2009.

28. Su Y, Lee CN, Cheng WF, Shau WY, Chow SN, Hsieh FJ. Decreased Maternal Serum Placenta Growth Factor in Early Second Trimester and Preeclampsia. Obstetrics & Gynecology. 2001;97:898–904.

29. Tidwell S, Ho HN, Chiu WH, Torry RJ, Torry DS. Low maternal serum levels of placenta growth factor as an antecedent of clinical preeclampsia. American journal of obstetrics and gynecology. 2001;184 6:1267–72.

30. Agrawal S, Shinar S, Cerdeira AS, Redman C, Vatish M. Predictive Performance of PlGF (Placental Growth Factor) for Screening Preeclampsia in Asymptomatic Women. Hypertension. 2019;74(5):1124–1135.

31. MacDonald TM, Tran CH, Kaitu’u-Lino TJ, et al. Assessing the sensitivity of placental growth factor and soluble fms-like tyrosine kinase 1 at 36 weeks’ gestation to predict small-for-gestational-age infants or late-onset preeclampsia: a prospective nested case-control study. BMC Pregnancy and Childbirth. 2018;18.

32. Levine RJ, Lam C, Qian C, et al. Soluble endoglin and other circulating antiangiogenic factors in preeclampsia. The New England journal of medicine. 2006;355 10:992–1005.

33. Zhang J, Klebanoff MA, Roberts JMD. Prediction of Adverse Outcomes by Common Definitions of Hypertension in Pregnancy. Obstetrics & Gynecology. 2001;97:261–267.

34. Roberts JM. Preeclampsia: new approaches but the same old problems. Am J Obstet Gynecol. 2008;199:443–444.

35. Serra B, Mendoza M, Scazzocchio E, et al. A new model for screening for early-onset preeclampsia. American Journal of Obstetrics & Gynecology. 2020;222:608.e1-608.e18.

36. Rana S, Lemoine E, Granger JP, Ananth KS. Preeclampsia. Circulation Research. 2019;124(7):1094–1112.

37. Baniecki H, Kretowicz W, Piatyszek P, Wisniewski J, Biecek P. dalex: Responsible Machine Learning with Interactive Explainability and Fairness in Python. Journal of Machine Learning Research. 2021;22:1–7.

38. Leaños-Miranda A, Navarro-Romero CS, Sillas-Pardo LJ, Ramírez-Valenzuela KL, Isordia-Salas I, Jiménez-Trejo LM. Soluble Endoglin As a Marker for Preeclampsia, Its Severity, and the Occurrence of Adverse Outcomes. Hypertension. 2019.

