## Appendix for "Preeclampsia Predictor with Machine Learning: A Comprehensive and Bias-Free Machine Learning Pipeline"

**Fairness Metrics**

To determine the fairness of our model, we identified and calculated a set of metric ratios as discussed below to determine the subgroup(s) for which the threshold of the four-fifths rule was violated. We then plotted a *ceteris paribus* cutoff plot for the subgroup with bias and adjusted the classification threshold accordingly. Fairness metrics create a framework for measuring discrimination based on the characteristics attributes. In order to better understand fairness metrics, we assume a source distribution (Y, X, A), where Y is the target label, X is the set of available features and $A \in\{0, 1\}$ is the characteristics attribute. For the sake of simplicity, we define A as a binary attribute, but A can have many labels, such as race. A predictor of Y is defined as $\hat{Y}=f\left( X,A \right)$. For a model to be deemed fair, it should be the case that f does not discriminate with respect to A. ^15^ In a practical setting, ratios of fairness metrics are calculated for protected groups and the privilege group. A model is considered unfair when such a ratio crosses a certain threshold. Typically, the Four-Fifths rule, which states that if the selection rate for a protected group is less than 80 percent of that of the group with the highest selection rate, there is an adverse impact on that group, is used as a threshold. Supplement Table 9 summarizes the fairness metrics and their respective mapping to evaluation metrics used in our analysis. The fairness metrics have been shown to be effective in the following situations: Predictive Parity (PP) is well-suited for predicting the likelihood of recidivism among high-risk oﬀenders; Accuracy Equality (AE) assures good credit scores are assigned to people with actual good credit and bad credit scores are assigned to people with actual bad credit for their group and finally Statistical Parity (SP) is appropriate for situations related to admission or employment, where an organization might prefer to have equal proportion of acceptance across different genders or racial backgrounds.

In our work, the EOR and PPR are the primary fairness metrics of interest. Unlike demographic parity, $Pr\{ \hat{Y} = 1 | A = 0\} = Pr\{ \hat{Y} = 1 | A = 1\} ,$ a widely used measure for fairness that ensures that there are no correlation between a decision made by predictor $\hat{Y}$ and characteristics attribute A, EOR and PER allow $\hat{Y}$ to be dependent on A, but only through the target variable Y. Thus, both metrics allow the feature to directly predict Y, but prohibits abusing A as a proxy for Y. EOR requires $\hat{Y}$ to have equal true positive rate across the two subgroups, $A=1$ and $A=0$: $Pr\{ \hat{Y} = 1 | A = 0, Y = 1 \} = Pr\{ \hat{Y} = 1 | A = 1, Y = 1 \}$. PER requires $\hat{Y}$ to have equal false positive rate across the two subgroups $A=1$ and $A=0$: $Pr\{\hat{Y} = 1 | A = 0, Y = 0 \} = Pr\{\hat{Y} = 1 | A = 1, Y = 0 \}$.

The Fairness evaluation is conducted using the python package Dalex: Responsible Machine Learning in Python. ^37^ Predictions for multiple trials were aggregated to produce a fairness metric ratio. This is calculated by taking the ratio of the respective evaluation metrics. In our analysis, the focus is on the characteristics attribute of race. For further evaluation, we will denote $A\in\{\text{Asian, Black, Hispanic, Other, White}\}$ as the race attribute. We denote the Non-Hispanic White participants ($A=White$) as the privileged group, while all other races are considered as protected group. For i$\in A \setminus\{ White \}$, given $TPR_{i}$ is the true positive rate of race $i$ and $TPR_{White}$ is the true positive rate of privileged subgroup, the equal opportunity ratio of a race $i$ ($EOR_{i}$) is calculated by: $EOR_{i}=\frac{TPR_{i}}{TPR_{White}}$ and the predictive equality ratio of race $i$ ($PER_{i}$) is calculated by: $PER_{i}=\frac{FPR_{i}}{FPR_{White}}$

***Ceteris Paribus* Cutoff Plot**

Consider the situation where we want not just a measure of how biased our model is for a certain race, but a measure that summarizes the bias across different races. This can be done through calculating the parity loss. Parity loss of EOR can be calculated using the following formulation:

$$Parity loss of EOR =\sum_{i \in A \setminus\{ White \}} \left| log \frac{{TPR}_{i}}{{TPR}_{Whjte}} \right|=\sum_{i \in A \setminus\{ White \}} \left| log {EOR}_{i} \right|$$

Similarly, the parity loss of PER can be calculated by:

$$Parity loss of PER =\sum_{i \in A \setminus\{ White \}} \left| log \frac{{FPR}_{i}}{{FPR}_{Whjte}} \right|=\sum_{i \in A \setminus\{ White \}} \left| log {PER}_{i} \right|$$

In order to mitigate the bias identified through parity loss, a post-processing algorithm, *ceteris paribus* cutoff plotting was used. This is a model-agnostic algorithm, which works for models with different structures, such as neural networks, random forests, boosting models and linear models. ^37^ Similar to the process of constructing a ROC, ceteris paribus cutoff plotting works by directly altering the classification threshold, but instead of visualizing the impact on TPR and FPR, the parity loss is used, and alteration of the classification threshold is only performed on a specific subgroup. Typically, we select the threshold by identifying the value that minimizes the parity loss.

**Software packages**

We developed our pipeline in Python 3. Instructions about how to run the experiments are provided in the Github repository. We also conducted the bias mitigation experiments using Dalex. PDPs were generated using the PDPbox package. Dataset balancing was done using the imbalanced-learn package. The model used for generating our results was trained using the XGBoost package. Link to github respitory:

https://github.com/adamcatto/NICHD_NuMom2b_Data_Challenge
