## Supplement Tables for "Preeclampsia Predictor with Machine Learning: A Comprehensive and Bias-Free Machine Learning Pipeline"

**Supplement Table 1:** Study population characteristic for demographic, family history and diet intake features

| Parameter | sPE + E | Early sPE | Late sPE + E | NPH | sPE+ E vs NPH p value* | Early sPE vs Late sPE + E p value* |
| --- | --- | --- | --- | --- | --- | --- |
| Demographics |  |  |  |  |  |  |
| No. of cases | 278 | 58 | 220 | 1480 | -- | -- |
| Maternal Age | 26 (22-31) | 25 (21-29) | 27 (22-32) | 27 (22-31) | 0.3 | 0.02 |
| Maternal Age |  |  |  |  |  |  |
| 13-17 | 9 (3.2) | 3 (5.2) | 6 (2,7) | 37 (2.5) | -- | -- |
| 18-34 | 236 (84.9) | 52 (89.7) | 184 (83.6) | 1311 (88.6) |  |  |
| 35-39 | 27 (9.7) | 3 (5.2) | 24 (10.9) | 115 (7.8) | -- | -- |
| $\geq40$ | 6 (2.2) | 0 (0.0) | 6 (2.7) | 16 (1.0) | -- | -- |
| Race |  |  |  |  | 0.03 | 0.67 |
| Non-Hispanic White | 144 (51.8) | 30 (51.7) | 114 (51.8) | 882 (59.6) | -- | -- |
| Non-Hispanic Black | 58 (20.9) | 8 (13.7) | 50 (22.7) | 204 (13.8) | -- | -- |
| Hispanic | 48 (17.3) | 14 (24.1) | 34 (15.5) | 263 (17.8) | -- | -- |
| Asian | 10 (3.6) | 3 (5.2) | 7 (3.2) | 50 (3.4) | -- | -- |
| Other | 18 (6.5) | 3 (5.2) | 15 (6.8) | 80 (5.4) | -- | -- |
| Parity | 1 (1-2) | 1 (1-1) | 1 (1-2) | 1 (1-2) | 0.243 | 0.132 |
| Family History |  |  |  |  |  |  |
| Preeclampsia, eclampsia, and |  |  |  |  |  |  |
| hypertension in close relatives | 37 (13.3) | 6 (10.3) | 31 (13.5) | 152 (10.3) | 0.17 | 0.52 |
| Diet intake |  |  |  |  |  |  |
| Cholesterol | 206 (140-291) | 221 (175-271) | 198 (132-295) | 179 (125-258) | 0.002 | 0.15 |
| Saturated fat | 20 (15-28) | 22 (16-32) | 20 (15-27) | 19 (14-26) | 0.006 | 0.14 |
| Monounsaturated fatty acids | 25 (18-34) | 28 (18-38) | 25 (18-33) | 23 (17-31) | 0.009 | 0.17 |
| Polyunsaturated fatty acids | 14 (10-18) | 14 (11-18) | 14 (9-17) | 12 (9-17) | 0.009 | 0.23 |

*** p values calculated with Welch’s t-test, Mann–Whitney U test, Chi Square test, or Fisher Exact test where appropriate.**

**Supplement Table 2:** Study population characteristic for ultrasound feature

| **Parameter** | **sPE + E** | | | **NPH** | | | **sPE + E vs NPH p value*** | | |
| --- | --- | --- | --- | --- | --- | --- | --- | --- | --- |
|  | V1 | V2 | V3 | V1 | V2 | V3 | V1 | V2 | V3 |
| Adrenal Gland Meas. |  |  |  |  |  |  |  |  |  |
| Fetal Zone Length | -- | -- | 11 (9-14) | -- | -- | 12 (10-14) | -- | -- | 0.11 |
| Fetal cardiac activity |  |  |  |  |  |  |  |  |  |
| Abdominal Circumference | -- | 14 (13-16) | 24 (22-25) | -- | 14 (13-15) | 23 (22-25) | -- | 0.453 | 0.027 |
| Fetal Biometery |  |  |  |  |  |  |  |  |  |
| Disclosable condition* | -- | 3 (1.1) | 6 (2.2) | -- | 53 (3.6) | 49 (3.3) | -- | 0.02 | 0.35 |
| Right Uterine Artery |  |  |  |  |  |  |  |  |  |
| Early Diastolic Notch | 17 (6) | 104 (37) | 64 (23) | 131 (9) | 339 (23) | 142 (10) | 0.7 | $<.001$ | $<.001$ |
| Resistance Index | .75 (.68-.81) | .59 (.49-.71) | .57 (.48-.68) | .69 (.58-.81) | .55 (.47-.64) | .49 (.43-.57) | 0.05 | $<.001$ | $<.001$ |
| Pulsatility Index | 1.7 (1.3-2.1) | 1 (.76-1.5) | .98 (.71-1.28) | 1.4 (.99-2.1) | .9 (.7-1.2) | .74 (.61-.95) | 0.04 | $<.001$ | $<.001$ |
| Systolic/Diastolic Ratio | 4 (3.2-5.8) | 2.5 (2-3.5) | 2.3 (1.9-3.1) | 3.3 (2.4-5.3) | 2.2 (1.9-2.8) | 2 (1.8-2.3) | 0.023 | $<.001$ | $<.001$ |
| Peak Systolic Velocity | 52 (41-67) | 75 (53-98) | 87 (64-115) | 55 (38-74) | 80 (56-110) | 92 (61-126) | 0.4 | 0.02 | 0.1 |
| Left Uterine Artery |  |  |  |  |  |  |  |  |  |
| Early Diastolic Notch | 20 (7) | 121 (44) | 71 (26) | 140 (10) | 393 (27) | 177 (12) | 0.4 | <0.001 | <0.001 |
| Resistance Index | .79 (.69-.85) | .63 (.53-0.73) | .58 (.50-.68) | .74 (.63-.81) | .57 (.49-.66) | .50 (.45-.59) | 0.019 | <0.001 | <0.001 |
| Pulsatility Index | 2 (1.4-2.4) | 1.2 (0.9-1.6) | .88 (.75-1.2) | 1.6 (1.2-2.1) | .95 (.75-1.3) | .78 (.64-1) | 0.02 | <.001 | <.001 |
| Systolic/Diastolic Ratio | 4.6 (3.1-5.9) | 2.7 (2.1-3.7) | 2.4 (2-3.1) | 3.8 (2.7-5.2) | 2.3 (1.9-2.9) | 2 (1.8-2.4) | 0.1 | <0.001 | <0.001 |
| Peak Systolic Velocity | 52 (44-69) | 81 (58-115) | 96 (66-129) | 58 (41-81) | 91 (60-119) | 108 (72-144) | 0.334 | 0.018 | 0.012 |
| **Parameter** | **Early sPE** |  |  | **Late sPE + E** |  |  | **Early sPE vs Late sPE + E p value** | | |
|  | V1 | V2 | V3 | V1 | V2 | V3 | V1 | V2 | V3 |
| Adrenal Gland Meas. |  |  |  |  |  |  |  |  |  |
| Fetal Zone Length | -- | -- | 9 (8-11) | -- | -- | 11 (9-14) | -- | -- | 0.07 |
| Fetal cardiac activity |  |  |  |  |  |  |  |  |  |
| Abdominal Circumference | -- | 14 (13-15) | 22 (20-24) | -- | 14 (13-16) | 24 (22-25) | -- | 0.008 | <0.001 |
| Fetal Biometery |  |  |  |  |  |  |  |  |  |
| Disclosable condition^1^ | -- | 0 (0.0) | 4 (6.9) | -- | 3 (1.3) | 2 (0.9) | -- | <0.01 | 1 |
| Right Uterine Artery |  |  |  |  |  |  |  |  |  |
| Early Diastolic Notch | 5 (9) | 32 (55) | 24 (41) | 12 (5) | 72 (31) | 40 (18) | 1 | <.001 | <.001 |
| Resistance Index | .77 (.72-.84) | .69 (.59-.77) | .75 (.63-.79) | .75 (.68-.81) | .57 (.49-.68) | .55 (.48-.61) | 0.3 | <.001 | <.001 |
| Pulsatility Index | 1.9 (1.5-2.3) | 1.5 (1-1.9) | 1.7 (1.2-2.2) | 1.7 (1.3-2) | .95 (.73-1.3) | .89 (.7-1.1) | 0.289 | <.001 | <.001 |
| Systolic/Diastolic Ratio | 4.8 (3.8-6.8) | 3.3 (2.5-4.3) | 4 (2.7-4.8) | 4 (3.1-5.1) | 2.3 (1.9-3.1) | 2.1 (1.9-2.6) | 0.17 | <.001 | <.001 |
| Peak Systolic Velocity | 52 (45-65) | 63 (46-86) | 75 (64-94) | 52 (41-67) | 79 (55-102) | 93 (64-116) | 0.3 | 0.002 | 0.07 |
| Left Uterine Artery |  |  |  |  |  |  |  |  |  |
| Early Diastolic Notch | 7 (12) | 33 (57) | 23 (40) | 13 (6) | 88 (38) | 48 (21) | 1 | 0.001 | 0.002 |
| Resistance Index | .83 (.78-.85) | .69 (.63-.76) | .66 (.62-.70) | .76 (.66-.82) | .61 (.52-.72) | .54 (.49-.64) | 0.038 | <0.001 | <0.001 |
| Pulsatility Index | 5.3 (4.3-6.4) | 3.3 (2.8-4.2) | 3 (2.7-3.4) | 4 (2.9-5.5) | 2.55 (2-3.5) | 2.2 (1.9-2.8) | 0.05 | <.001 | <.001 |
| Systolic/Diastolic Ratio | 1 (.8-1.4) | 2.1 (2.1-2.4) | 1.4 (1.2-1.9) | 1.4 (1.1-1.6) | 1.8 (1.3-2.2) | 1.1 (.8-1.5) | 0.1 | <0.001 | <0.001 |
| Peak Systolic Velocity | 48 (35-63) | 74 (55-95) | 91 (65-109) | 58 (46-72) | 82 (59-117) | 96 (69-131) | 0.13 | 0.057 | 0.266 |

**^1^** Disclosable condition include: a) fetal demise, b) estimated fetal weight $<5$th percentile, c) incidental finding of oligohydramnios, d) obvious fetal bradycardia or tachycardia, e) incidental detection of complete or partial placenta previa or vasa previa, d) incidental detection of major fetal structural malformation or hydrops.

*** p values calculated with Welch’s t-test, Mann–Whitney U test, Chi Square test, or Fisher Exact test where appropriate.**

**Supplement Table 3:** Study population characteristic for placental analytes features

| **Parameter** | **sPE + E** | | **NPH** | | **sPE+ E vs NPH p value*** | |
| --- | --- | --- | --- | --- | --- | --- |
|  | V1 | V2 | V1 | V2 | V1 | V2 |
| Endoglin | 6.4 (5.3-7.7) | 5.8 (5-6.8) | 6.3 (5.4-7.3) | 5.4 (4.7-6.2) | 0.129 | <0.001 |
| Inhibin A | 333 (238-460) | 225 (176-316) | 310 (225-425) | 200 (156-266) | 0.03 | <0.001 |
| ADAM12 | 4.5 (3.4-5.7) | 10 (7.8-12.3) | 4.7 (3.7-6) | 9.9 (7.8-12.2) | 0.03 | <0.001 |
| SFLT1 | 816 (627-1120) | 901 (628-1190) | 910 (692-1190) | 882 (638-1216) | 0.001 | 0.282 |
| VEGF | 0.98 (0.69-2.02) | 1.10 (0.76-1.88) | 0.86 (0.65-1.28) | 0.99 (0.73-1.58) | <0.001 | 0.034 |
| AFP | 13.34 (7.86-19.50) | 46.2 (36.5-63.4) | 13.8 (8.3-20.3) | 46.9 (35.4-60.3) | 0.171 | 0.348 |
| fbHCG | 19.5 (12.7-29.6) | 4 (2.6-6.5) | 21.1 (13.9-32.1) | 4.1 (2.7-6.5) | 0.029 | 0.469 |
| PAPP-A | 787 (357-1594) | 9017 (4486-15896) | 988 (477-1849) | 9242 (5527-14857) | 0.003 | 0.206 |
| PlGF | 36 (25-49) | 148 (90-239) | 42 (29-58) | 198 (134-291) | <0.001 | <0.001 |
| sFLT1-to-PLGF ratio | 21.8 (15.7-33.5) | 5.4 (3.8-9.3) | 21.2 (14.2-31.6) | 4.5 (3-6.6) | 0.069 | <0.001 |
| **Parameter** | **Early sPE** | | **Late sPE + E** | | **Early sPE vs Late sPE + E p value** | |
|  | V1 | V2 | V1 | V2 | V1 | V2 |
| Endoglin | 6.2 (5.4-7.5) | 6.3 (5.4-7.3) | 6.4 (5.3-7.7) | 5.7 (4.9-6.6) | 0.361 | 0.014 |
| Inhibin A | 340 (248-484) | 307 (209-441) | 332 (235-459) | 215 (169-279) | 0.358 | <0.001 |
| ADAM12 | 3.8 (3-5) | 10.2 (8.2-12.9) | 4.6 (3.5-6) | 10 (7.8-12.3) | 0.358 | <0.001 |
| SFLT1 | 787 (545-947) | 936 (655-1221) | 856 (655-1149) | 885 (592-1179) | 0.005 | 0.166 |
| VEGF | 1.16 (0.74-2.65) | 0.88 (0.66-1.31) | 0.96 (0.69-1.87) | 1.15 (0.76-1.94) | 0.27 | 0.041 |
| AFP | 13.2 (8.3-18.5) | 49.4 (36.6-77) | 13.4 (7.8-19.7) | 45.4 (36-61.7) | 0.278 | 0.047 |
| fbHCG | 20.6 (13.8-30.3) | 4.9 (3.3-8.3) | 19.3 (12.1-29) | 3.9 (2.5-6) | 0.184 | 0.006 |
| PAPP-A | 561 (231-884) | 7386 (4339-13758) | 877 (414-1838) | 9526 (4613-16075) | <0.001 | 0.076 |
| PlGF | 26 (21-36) | 82 (51-126) | 39 (27-53) | 165 (112-256) | <0.001 | <0.001 |
| sFLT1-to-PLGF ratio | 26.1 (20-37.5) | 9.8 (5.7-17.1) | 21.2 (15.4-32.9) | 4.5 (3.4-8) | 0.024 | <0.001 |

*** p values calculated with Welch’s t-test, Mann–Whitney U test, Chi Square test, or Fisher Exact test where appropriate.**

**Supplement Table 4:** Study population characteristic for physiology and sleep features

| **Parameter** | **sPE + E** | | | **NPH** | | | **sPE + E vs NPH p value** | | |
| --- | --- | --- | --- | --- | --- | --- | --- | --- | --- |
|  | V1 | V2 | V3 | V1 | V2 | V3 | V1 | V2 | V3 |
| Systolic Blood Pressure | 112 (106-120) | 114 (108-120) | 118 (110-124) | 108 (100-115) | 110 (100-116) | 110 (102-115) | <0.001 | <0.001 | <0.001 |
| 1 visit % CHG | -- | 0 (-7-7) | 2 (-4-9) | -- | 0 (-6-8) | 0 (-6-7) | -- | 0.184 | 0.002 |
| 2 visit % CHG | -- | -- | 2 (-5-9) | -- | -- | 0 (-6-9) | -- | -- | 0.059 |
| Diastolic Blood Pressure | 70 (64-75) | 68 (62-76) | 70 (64-78) | 66 (60-70) | 64 (60-70) | 64 (60-70) | <0.001 | <0.001 | <0.001 |
| 1 visit % CHG | -- | -1 (-11-8) | 3 (-6-13) | -- | 0 (-10-7) | 0 (-7-10) | -- | 0.401 | 0.006 |
| 2 visit % CHG | -- | -- | 0 (-9-11) | -- | -- | 0 (-10-9) | -- | -- | 0.014 |
| Mean Arterial Pressure | 84 (79-90) | 83 (77-90) | 86 (80-92) | 80 (74-85) | 79 (74-84) | 80 (75-85) | <0.001 | <0.001 | <0.001 |
| BMI | 27 (23-32) | -- | -- | 24 (22-28) | -- | -- | <0.001 | -- | -- |
| Weight (LB) | 155 (135-190) | 160 (142-196) | 172 (149-207) | 144 (128-166) | 149 (133-173) | 160 (143-184) | <0.001 | <0.001 | <0.001 |
| 1 visit % CHG | -- | 3 (1-6) | 7 (4-10) | -- | 4 (2-6) | 7 (4-9) | -- | 0.038 | 0.284 |
| 2 visit % CHG | -- | -- | 10 (7-15) | -- | -- | 11 (7-15) | -- | -- | 0.395 |
| Waist Circumference | 97 (89-108) | -- | -- | 91 (84-100) | -- | -- | <0.001 | -- | -- |
| Neck Circumference | 34 (32-36) | -- | -- | 32 (31-34) | -- | -- | <0.001 | -- | -- |
| Sleep Apnea | 2 (0.7) | -- | 2 (0.7) | 12 (0.8) | -- | 12 (0.8) | 0.717 | -- | 1 |
| **Parameter** | **Early sPE** | | | **Late sPE + E** | | | **Early sPE vs Late sPE + E p value** | | |
|  | V1 | V2 | V3 | V1 | V2 | V3 | V1 | V2 | V3 |
| Systolic Blood Pressure | 113 (110-122) | 116 (110-122) | 120 (112-127) | 112 (106-120) | 112 (108-120) | 116 (110-122) | 0.202 | 0.042 | 0.002 |
| 1 visit % CHG | -- | 2 (-5-6) | 4 (0-11) | -- | 0 (-8-7) | 2 (-4-8) | -- | 0.179 | 0.044 |
| 2 visit % CHG | -- | -- | 5 (0-12) | -- | -- | 0 (-6-9) | -- | -- | 0.009 |
| Diastolic Blood Pressure | 70 (66-78) | 70 (66-78) | 78 (68-84) | 70 (62-74) | 68 (60-74) | 70 (62-76) | 0.112 | 0.007 | <0.001 |
| 1 visit % CHG | -- | 0 (-4-9) | 7 (-3-19) | -- | -2 (-11-6) | 3 (-6-12) | -- | 0.068 | 0.045 |
| 2 visit % CHG | -- | -- | 10 (-4-17) | -- | -- | 0 (-9-9) | -- | -- | 0.006 |
| Mean Arterial Pressure | 85 (80-92) | 87 (80-91) | 90 (84-97) | 84 (78-89) | 83 (77-89) | 85 (79-91) | 0.103 | 0.008 | <0.001 |
| BMI | 29 (25-33) | -- | -- | 27 (23-32) | -- | -- | 0.166 | -- | -- |
| Weight (LB) | 164 (137-196) | 167 (137-196) | 163 (143-189) | 152 (135-187) | 160 (143-196) | 174 (151-209) | 0.315 | 0.322 | 0.057 |
| 1 visit % CHG | -- | 2 (1-4) | 6 (3-8) | -- | 4 (1-6) | 7 (4-10) | -- | 0.002 | 0.028 |
| 2 visit % CHG | -- | -- | 8 (4-14) | -- | -- | 11 (8-15) | -- | -- | 0.003 |
| Waist Circumference | 100 (90-110) | -- | -- | 96 (89-107) | -- | -- | 0.128 | -- | -- |
| Neck Circumference | 34 (32-36) | -- | -- | 34 (32-35) | -- | -- | 0.097 | -- | -- |
| Sleep Apnea | 0 (0.0) | -- | 0 (0.0) | 3 (1.3) | -- | 2 (0.9) | 1 | -- | 1 |

*** p values calculated with Welch’s t-test, Mann–Whitney U test, Chi Square test, or Fisher Exact test where appropriate.**

**Supplement Table 5:** Study population characteristic for medical conditions features

| **Parameter** | **sPE + E** | | | | **NPH** | | | | **sPE + E vs NPH p value** | | | |
| --- | --- | --- | --- | --- | --- | --- | --- | --- | --- | --- | --- | --- |
|  | V1 | V2 | V3 | V4 | V1 | V2 | V3 | V4 | V1 | V2 | V3 | V4 |
| Liver/gall bladder disease | 6 (2.2) | 5 (1.8) | 4 (1.4) | 2 (0.7) | 42 (2.8) | 38 (2.6) | 35 (2.4) | 22 (1.5) | 0.688 | 0.533 | 0.383 | 0.409 |
| Diabetes |  |  |  |  |  |  |  |  |  |  |  |  |
| Gestational | -- | 14 (5.0) | 5 (1.8) | 7 (2.5) | -- | 20 (1.4) | 23 (1.6) | 29 (2.0) | -- | <0.001 | 0.138 | 0.494 |
| Excluding gestational | 13 (4.7) | 14 (5.0) | 14 (5.0) | 13 (4.7) | 21 (1.4) | 20 (1.4) | 36 (2.4) | 47 (3.2) | 0.001 | <0.001 | 0.032 | 0.209 |
| Lupus | 1 (0.4) | 1 (0.4) | 1 (0.4) | 1 (0.4) | 7 (0.5) | 6 (0.4) | 5 (0.3) | 7 (0.5) | 1 | 1 | 1 | 1 |
| PCOS | 13 (4.7) | 14 (5.0) | 13 (4.7) | 8 (2.9) | 63 (4.3) | 53 (3.6) | 46 (3.1) | 45 (3.0) | 0.746 | 0.402 | 0.28 | 1 |
| Blood clots | 5 (1.8) | 4 (1.4) | 3 (1.1) | 3 (1.1) | 16 (1.1) | 11 (0.7) | 10 (0.7) | 11 (0.7) | 0.358 | 0.298 | 0.459 | 0.476 |
| Hypertension | 10 (3.6) | 10 (3.6) | 15 (5.4) | 74 (27) | 32 (2.2) | 25 (1.7) | 23 (1.6) | 17 (1.1) | 0.137 | 0.068 | <0.001 | <0.001 |
| Cardiovascular Abnormalities |  |  |  |  |  |  |  |  |  |  |  |  |
| Valvular heart disease | 1 (0.4) | 1 (0.4) | 1 (0.4) | 1 (0.4) | 16 (1.1) | 12 (0.8) | 11 (0.7) | 11 (0.7) | 0.498 | 0.708 | 0.703 | 0.704 |
| Other structural heart disease | 2 (0.7) | 2 (0.7) | 3 (1.1) | 0 (0.0) | 15 (1.0) | 12 (0.8) | 11 (0.7) | 4 (0.3) | 1 | 1 | 0.714 | 1 |
| Cardiac arrhythmias | 4 (1.4) | 5 (1.8) | 5 (1.8) | 4 (1.4) | 33 (2.2) | 26 (1.8) | 33 (2.2) | 24 (1.6) | 0.501 | 1 | 0.824 | 1 |
| Kidney disease | 7 (2.5) | 9 (3.2) | 9 (3.2) | 10 (3.6) | 31 (2.1) | 28 (1.9) | 22 (1.5) | 28 (1.9) | 0.651 | 0.262 | 0.081 | 0.111 |
| Thrombocytopenia | 1 (0.4) | 1 (0.4) | 3 (1.1) | 7 (2.5) | 13 (0.9) | 14 (0.9) | 16 (1.1) | 19 (1.3) | 0.71 | 0.489 | 1 | 0.169 |
| Anemia | 28 (10) | 30 (11) | 32 (12) | 31 (11) | 168 (11) | 150 (10) | 162 (11) | 169 (11) | 0.676 | 1 | 1 | 1 |
| Rheumatoid arthritis | 2 (0.7) | 2 (0.7) | 1 (0.4) | 1 (0.4) | 16 (1.1) | 15 (1.0) | 12 (0.8) | 6 (0.4) | 1 | 0.753 | 0.706 | 1 |
| Complete blood count |  |  |  |  |  |  |  |  |  |  |  |  |
| Hemoglobin g/dL | -- | -- | 13 (12-14) | -- | -- | -- | 13 (12-14) | -- | -- | -- | 0.122 | -- |
| Hematocrit % | -- | -- | 38 (36-40) | -- | -- | -- | 38 (36-40) | -- | -- | -- | 0.337 | -- |
| MCV fL/cell | -- | -- | 89 (85-92) | -- | -- | -- | 90 (87-93) | -- | -- | -- | 0.003 | -- |
| Platelet count x10 3 /mm 3 | -- | -- | 247 (211-299) | -- | -- | -- | 247 (212-285) | -- | -- | -- | 0.208 | -- |
| **Parameter** | **Early sPE** | | | | **Late sPE + E** | | | | **Early sPE vs Late sPE + E p value** | | | |
|  | V1 | V2 | V3 | V4 | V1 | V2 | V3 | V4 | V1 | V2 | V3 | V4 |
| Liver/gall bladder disease | 1 (1.7) | 1 (1.7) | 1 (1.7) | 1 (1.7) | 5 (2.2) | 4 (1.7) | 3 (1.3) | 1 (0.4) | 1 | 1 | 0.523 | 0.374 |
| Diabetes |  |  |  |  |  |  |  |  |  |  |  |  |
| Gestational | -- | 3 (5.2) | 1 (1.7) | 1 (1.7) | -- | 11 (4.8) | 4 (1.7) | 6 (2.6) | -- | 1 | 1 | 1 |
| Excluding gestational | 3 (5.2) | 3 (5.2) | 2 (3.4) | 3 (5.2) | 10 (4.4) | 11 (4.8) | 12 (5.2) | 10 (4.4) | 0.738 | 1 | 1 | 0.738 |
| Lupus | 1 (1.7) | 1 (1.7) | 1 (1.7) | 1 (1.7) | 0 (0.0) | 0 (0.0) | 0 (0.0) | 0 (0.0) | 0.209 | 0.211 | 0.168 | 0.209 |
| PCOS | 1 (1.7) | 1 (1.7) | 0 (0.0) | 0 (0.0) | 12 (5.2) | 13 (5.7) | 13 (5.7) | 8 (3.5) | 0.313 | 0.313 | 0.133 | 0.211 |
| Blood clots | 1 (1.7) | 1 (1.7) | 1 (1.7) | 0 (0.0) | 4 (1.7) | 3 (1.3) | 2 (0.9) | 3 (1.3) | 1 | 1 | 0.426 | 1 |
| Hypertension | 2 (3.4) | 2 (3.4) | 5 (8.6) | 19 (33) | 8 (3.5) | 8 (3.5) | 10 (4.4) | 55 (24) | 1 | 1 | 0.143 | 0.245 |
| Cardiovascular Abnormalities |  |  |  |  |  |  |  |  |  |  |  |  |
| Valvular heart disease | 0 (0.0) | 0 (0.0) | 0 (0.0) | 1 (1.7) | 1 (0.4) | 1 (0.4) | 1 (0.4) | 0 (0.0) | 1 | 1 | 1 | 0.209 |
| Other structural heart disease | 1 (1.7) | 1 (1.7) | 1 (1.7) | 0 (0.0) | 1 (0.4) | 1 (0.4) | 2 (0.9) | 0 (0.0) | 0.375 | 0.378 | 0.426 | 1 |
| Cardiac arrhythmias | 0 (0.0) | 0 (0.0) | 0 (0.0) | 0 (0.0) | 4 (1.7) | 5 (2.2) | 5 (2.2) | 4 (1.7) | 0.583 | 0.587 | 0.593 | 0.583 |
| Kidney disease | 6 (10.3) | 7 (12.1) | 4 (6.9) | 7 (12.1) | 1 (0.4) | 2 (0.9) | 5 (2.2) | 3 (1.3) | <0.001 | <0.001 | 0.046 | <0.001 |
| Thrombocytopenia | 0 (0.0) | 0 (0.0) | 1 (1.7) | 2 (3.4) | 1 (0.4) | 1 (0.4) | 2 (0.9) | 5 (2.2) | 1 | 1 | 0.426 | 0.639 |
| Anemia | 3 (5.2) | 6 (10) | 3 (5.2) | 6 (10) | 25 (11) | 24 (11) | 29 (12) | 25 (11) | 0.22 | 1 | 0.312 | 1 |
| Rheumatoid arthritis | 1 (1.7) | 1 (1.7) | 0 (0.0) | 1 (1.7) | 1 (0.4) | 1 (0.4) | 1 (0.4) | 0 (0.0) | 0.375 | 0.378 | 1 | 0.209 |
| Hemoglobin g/dL | -- | -- | 13 (12-14) | -- | -- | -- | 13 (12-14) | -- | -- | -- | 0.409 | -- |
| Hematocrit % | -- | -- | 38 (36-40) | -- | -- | -- | 38 (36-40) | -- | -- | -- | 0.359 | -- |
| MCV fL/cell | -- | -- | 88 (84-92) | -- | -- | -- | 89 (86-92) | -- | -- | -- | 0.098 | -- |
| Platelet count x10 3 /mm 3 | -- | -- | 275 (224-319) | -- | -- | -- | 243 (207-292) | -- | -- | -- | 0.011 | -- |

*** p values calculated with Welch’s t-test, Mann–Whitney U test, Chi Square test, or Fisher Exact test where appropriate.**

**Supplement Table 6:** Detail summary of early sPE vs NPH model performance per visit for RF.

| **Visits** | **AUC** | **Sensitivity (TP/ TP+FN)** | **Specificity (TN/ FP + TN)** | **PPV (TP/ TP +FP)** | **NPV (TN/FN+TN)** |
| --- | --- | --- | --- | --- | --- |
| **V1** | $\boldsymbol{0.73}\boldsymbol{\pm}\boldsymbol{0.11}$ | $\boldsymbol{0.71}\boldsymbol{\pm}\boldsymbol{0.14}$ | $\boldsymbol{0.63}\boldsymbol{\pm}\boldsymbol{0.14}$ | $\boldsymbol{0.66}\boldsymbol{\pm}\boldsymbol{0.10}$ | $\boldsymbol{0.70 \pm0.13}$ |
| **V2** | $\boldsymbol{0.87}\boldsymbol{\pm}\boldsymbol{0.07}$ | $\boldsymbol{0.86}\boldsymbol{\pm}\boldsymbol{0.10}$ | $\boldsymbol{0.68}\boldsymbol{\pm}\boldsymbol{0.13}$ | $\boldsymbol{0.74}\boldsymbol{\pm}\boldsymbol{0.09}$ | $\boldsymbol{0.84 \pm0.10}$ |
| **V3** | $\boldsymbol{0.89}\boldsymbol{\pm}\boldsymbol{0.07}$ | $\boldsymbol{0.88}\boldsymbol{\pm}\boldsymbol{0.10}$ | $\boldsymbol{0.72}\boldsymbol{\pm}\boldsymbol{0.15}$ | $\boldsymbol{0.77}\boldsymbol{\pm}\boldsymbol{0.10}$ | $\boldsymbol{0.86 \pm0.10}$ |
| **V4** | $\boldsymbol{0.94}\boldsymbol{\pm}\boldsymbol{0.04}$ | $\boldsymbol{0.92}\boldsymbol{\pm}\boldsymbol{0.07}$ | $\boldsymbol{0.78}\boldsymbol{\pm}\boldsymbol{0.12}$ | $\boldsymbol{0.81}\boldsymbol{\pm}\boldsymbol{0.08}$ | $\boldsymbol{0.91 \pm0.08}$ |

**Supplement Table 7:** Detail summary of late sPE+E vs NPH model performance per visit for RF

| **Visits** | **AUC** | **Sensitivity (TP/ TP+FN)** | **Specificity (TN/ FP + TN)** | **PPV (TP/ TP +FP)** | **NPV (TN/FN+TN)** |
| --- | --- | --- | --- | --- | --- |
| **V1** | $\boldsymbol{0.67}\boldsymbol{\pm}\boldsymbol{0.05}$ | $\boldsymbol{0.61}\boldsymbol{\pm}\boldsymbol{0.08}$ | $\boldsymbol{0.65}\boldsymbol{\pm}\boldsymbol{0.07}$ | $\boldsymbol{0.63}\boldsymbol{\pm}\boldsymbol{0.06}$ | $\boldsymbol{0.62}\boldsymbol{\pm}\boldsymbol{0.05}$ |
| **V2** | $\boldsymbol{0.69}\boldsymbol{\pm}\boldsymbol{0.05}$ | $\boldsymbol{0.60}\boldsymbol{\pm}\boldsymbol{0.08}$ | $\boldsymbol{0.68}\boldsymbol{\pm}\boldsymbol{0.07}$ | $\boldsymbol{0.65}\boldsymbol{\pm}\boldsymbol{0.05}$ | $\boldsymbol{0.63}\boldsymbol{\pm}\boldsymbol{0.05}$ |
| **V3** | $\boldsymbol{0.72}\boldsymbol{\pm}\boldsymbol{0.05}$ | $\boldsymbol{0.62}\boldsymbol{\pm}\boldsymbol{0.06}$ | $\boldsymbol{0.70}\boldsymbol{\pm}\boldsymbol{0.08}$ | $\boldsymbol{0.68}\boldsymbol{\pm}\boldsymbol{0.06}$ | $\boldsymbol{0.65}\boldsymbol{\pm}\boldsymbol{0.04}$ |
| **V4** | $\boldsymbol{0.81}\boldsymbol{\pm}\boldsymbol{0.04}$ | $\boldsymbol{0.69}\boldsymbol{\pm}\boldsymbol{0.07}$ | $\boldsymbol{0.77}\boldsymbol{\pm}\boldsymbol{0.07}$ | $\boldsymbol{0.75}\boldsymbol{\pm}\boldsymbol{0.06}$ | $\boldsymbol{0.72}\boldsymbol{\pm}\boldsymbol{0.05}$ |

**Supplement Table 8:** Detail summary of early sPE vs late sPE+E model performance per visit for 4 classifiers

| **Model** | **Visits** | **AUC** | **Sensitivity (TP/TP+FN)** | **Specificity (TN/FP+TN)** | **PPV (TP/TP+FP)** | **NPV (TN/FN+TN)** |
| --- | --- | --- | --- | --- | --- | --- |
| **LR** | **V1** | $\boldsymbol{0.61}\boldsymbol{\pm}\boldsymbol{0.13}$ | $\boldsymbol{0.55}\boldsymbol{\pm}\boldsymbol{0.15}$ | $\boldsymbol{0.60}\boldsymbol{\pm}\boldsymbol{0.16}$ | $\boldsymbol{0.59}\boldsymbol{\pm}\boldsymbol{0.12}$ | $\boldsymbol{0.57}\boldsymbol{\pm}\boldsymbol{0.10}$ |
|  | **V2** | $\boldsymbol{0.75}\boldsymbol{\pm}\boldsymbol{0.10}$ | $\boldsymbol{0.67}\boldsymbol{\pm}\boldsymbol{0.14}$ | $\boldsymbol{0.69}\boldsymbol{\pm}\boldsymbol{0.13}$ | $\boldsymbol{0.69}\boldsymbol{\pm}\boldsymbol{0.11}$ | $\boldsymbol{0.69}\boldsymbol{\pm}\boldsymbol{0.11}$ |
|  | **V3** | $\boldsymbol{0.80}\boldsymbol{\pm}\boldsymbol{0.09}$ | $\boldsymbol{0.72}\boldsymbol{\pm}\boldsymbol{0.13}$ | $\boldsymbol{0.75}\boldsymbol{\pm}\boldsymbol{0.14}$ | $\boldsymbol{0.75}\boldsymbol{\pm}\boldsymbol{0.11}$ | $\boldsymbol{0.73}\boldsymbol{\pm}\boldsymbol{0.10}$ |
|  | **V4** | $\boldsymbol{0.78}\boldsymbol{\pm}\boldsymbol{0.08}$ | $\boldsymbol{0.72}\boldsymbol{\pm}\boldsymbol{0.12}$ | $\boldsymbol{0.73}\boldsymbol{\pm}\boldsymbol{0.13}$ | $\boldsymbol{0.74}\boldsymbol{\pm}\boldsymbol{0.10}$ | $\boldsymbol{0.73}\boldsymbol{\pm}\boldsymbol{0.09}$ |
| **SVM** | **V1** | $\boldsymbol{0.60}\boldsymbol{\pm}\boldsymbol{0.11}$ | $\boldsymbol{0.48}\boldsymbol{\pm}\boldsymbol{0.17}$ | $\boldsymbol{0.64}\boldsymbol{\pm}\boldsymbol{0.17}$ | $\boldsymbol{0.58}\boldsymbol{\pm}\boldsymbol{0.13}$ | $\boldsymbol{0.57}\boldsymbol{\pm}\boldsymbol{0.10}$ |
|  | **V2** | $\boldsymbol{0.76}\boldsymbol{\pm}\boldsymbol{0.09}$ | $\boldsymbol{0.68}\boldsymbol{\pm}\boldsymbol{0.13}$ | $\boldsymbol{0.70}\boldsymbol{\pm}\boldsymbol{0.13}$ | $\boldsymbol{0.71}\boldsymbol{\pm}\boldsymbol{0.10}$ | $\boldsymbol{0.69}\boldsymbol{\pm}\boldsymbol{0.10}$ |
|  | **V3** | $\boldsymbol{0.81}\boldsymbol{\pm}\boldsymbol{0.08}$ | $\boldsymbol{0.72}\boldsymbol{\pm}\boldsymbol{0.12}$ | $\boldsymbol{0.74}\boldsymbol{\pm}\boldsymbol{0.13}$ | $\boldsymbol{0.74}\boldsymbol{\pm}\boldsymbol{0.10}$ | $\boldsymbol{0.73}\boldsymbol{\pm}\boldsymbol{0.09}$ |
|  | **V4** | $\boldsymbol{0.80}\boldsymbol{\pm}\boldsymbol{0.09}$ | $\boldsymbol{0.70}\boldsymbol{\pm}\boldsymbol{0.13}$ | $\boldsymbol{0.77}\boldsymbol{\pm}\boldsymbol{0.13}$ | $\boldsymbol{0.76}\boldsymbol{\pm}\boldsymbol{0.11}$ | $\boldsymbol{0.72}\boldsymbol{\pm}\boldsymbol{0.09}$ |
| **RF** | **V1** | $\boldsymbol{0.63}\boldsymbol{\pm}\boldsymbol{0.11}$ | $\boldsymbol{0.61}\boldsymbol{\pm}\boldsymbol{0.14}$ | $\boldsymbol{0.58}\boldsymbol{\pm}\boldsymbol{0.14}$ | $\boldsymbol{0.60}\boldsymbol{\pm}\boldsymbol{0.10}$ | $\boldsymbol{0.61}\boldsymbol{\pm}\boldsymbol{0.10}$ |
|  | **V2** | $\boldsymbol{0.79}\boldsymbol{\pm}\boldsymbol{0.11}$ | $\boldsymbol{0.70}\boldsymbol{\pm}\boldsymbol{0.16}$ | $\boldsymbol{0.75}\boldsymbol{\pm}\boldsymbol{0.13}$ | $\boldsymbol{0.75}\boldsymbol{\pm}\boldsymbol{0.12}$ | $\boldsymbol{0.72}\boldsymbol{\pm}\boldsymbol{0.12}$ |
|  | **V3** | $\boldsymbol{0.83}\boldsymbol{\pm}\boldsymbol{0.08}$ | $\boldsymbol{0.75}\boldsymbol{\pm}\boldsymbol{0.12}$ | $\boldsymbol{0.76}\boldsymbol{\pm}\boldsymbol{0.12}$ | $\boldsymbol{0.76}\boldsymbol{\pm}\boldsymbol{0.10}$ | $\boldsymbol{0.76}\boldsymbol{\pm}\boldsymbol{0.10}$ |
|  | **V4** | $\boldsymbol{0.84}\boldsymbol{\pm}\boldsymbol{0.09}$ | $\boldsymbol{0.73}\boldsymbol{\pm}\boldsymbol{0.13}$ | $\boldsymbol{0.77}\boldsymbol{\pm}\boldsymbol{0.13}$ | $\boldsymbol{0.77}\boldsymbol{\pm}\boldsymbol{0.10}$ | $\boldsymbol{0.75}\boldsymbol{\pm}\boldsymbol{0.10}$ |
| **XGBoost** | **V1** | $\boldsymbol{0.62}\boldsymbol{\pm}\boldsymbol{0.10}$ | $\boldsymbol{0.60}\boldsymbol{\pm}\boldsymbol{0.10}$ | $\boldsymbol{0.57}\boldsymbol{\pm}\boldsymbol{0.09}$ | $\boldsymbol{0.60}\boldsymbol{\pm}\boldsymbol{0.11}$ | $\boldsymbol{0.59}\boldsymbol{\pm}\boldsymbol{0.04}$ |
|  | **V2** | $\boldsymbol{0.76}\boldsymbol{\pm}\boldsymbol{0.09}$ | $\boldsymbol{0.43}\boldsymbol{\pm}\boldsymbol{0.13}$ | $\boldsymbol{0.92}\boldsymbol{\pm}\boldsymbol{0.08}$ | $\boldsymbol{0.85}\boldsymbol{\pm}\boldsymbol{0.13}$ | $\boldsymbol{0.62}\boldsymbol{\pm}\boldsymbol{0.06}$ |
|  | **V3** | $\boldsymbol{0.82}\boldsymbol{\pm}\boldsymbol{0.08}$ | $\boldsymbol{0.53}\boldsymbol{\pm}\boldsymbol{0.15}$ | $\boldsymbol{0.91}\boldsymbol{\pm}\boldsymbol{0.08}$ | $\boldsymbol{0.86}\boldsymbol{\pm}\boldsymbol{0.12}$ | $\boldsymbol{0.67}\boldsymbol{\pm}\boldsymbol{0.08}$ |
|  | **V4** | $\boldsymbol{0.82}\boldsymbol{\pm}\boldsymbol{0.09}$ | $\boldsymbol{0.52}\boldsymbol{\pm}\boldsymbol{0.14}$ | $\boldsymbol{0.93}\boldsymbol{\pm}\boldsymbol{0.07}$ | $\boldsymbol{0.88}\boldsymbol{\pm}\boldsymbol{0.11}$ | $\boldsymbol{0.67}\boldsymbol{\pm}\boldsymbol{0.07}$ |

**Supplement Table 9:** Fairness metrics and their respective mapping to evaluation metrics

| **Fairness Metrics** | **Evaluation Metrics** |
| --- | --- |
| Equal Opportunity Ratio (EOR ^15^ | True Positive Rate (TPR) |
| Predictive Parity Ratio (PPR) ^16^ | Positive Prediction Value (PPV) |
| Predictive Equality Ratio (PER) ^16^ | False Positive Rate (FPR) |
| Accuracy Equality Ratio (AER) ^17^ | Accuracy (ACC) |
| Statistical Parity Ratio (SPR) ^16^ | Positive Prediction Rate (PPR) |

**Supplement Table 10:** Abbreviations

| **Abbreviation** | **Meaning** |
| --- | --- |
| PE | Preeclampsia |
| sPE | Preeclampsia with severe features |
| E | Eclampsia |
| NPH | No pregnancy-related hypertension |
| RF | Random Forest |
| SVM | Support vector machines |
| XGBoost | eXtreme Gradient Boosting |
| LR | Logistic regression |
| AUC | Area under receiver operating characteristic curve |
| BMI | Body mass index |
| PlGF | Placental growth factor |
| sFLT1 | soluble Flt-1 |
| ADAM12 | A Disintegrin And Metalloprotease 12 |
| VEGF | Vascular endothelial growth factor |
| fbHCG | free beta-hCG |
| AFP | Alpha-Fetoprotein |
| **PDP** | **Partial Dependence Plot** |
| **SBP** | **Systolic blood pressure** |
| **DBP** | **Diastolic blood pressure** |
| **MAP** | **Mean Arterial Pressure** |
| **PPV** | **positive predictive value** |
| **HTN** | **Hypertension** |
| **EOR** | **Equal Opportunity Ratio** |
| **PPR** | **Predictive Parity Ratio** |
| **PER** | **Predictive Equality Ratio** |
| **AER** | **Accuracy Equality Ratio** |
| **SPR** | **Statistical Parity Ratio** |
