## Supplement Figure Legends for "Preeclampsia Predictor with Machine Learning: A Comprehensive and Bias-Free Machine Learning Pipeline"

**Supplement Figure 1:** Final study cohort selection process.

Out of the participants from the placental anlaytes sub-study we excluded participant with condition such as chronic hypertension, mild preeclampsia, and miss label for preeclampsia in order to focus on the participant that are most at risk.

**Supplement Figure 2:** Preeclamspia diagnosis flowchart according to nuMoM2b criteria.

nuMoM2b criteria is selected as the study outcome for our analysis.

**Supplement Figure 3:** sPE+E vs NPH feature importance for V1.

Features with top 30 feature importance values for RF are shown. For each feature, feature importance for both RF and XGBoost are both reported. Features from different visits are encoded in different colors.

**Supplement Figure 4:** sPE+E vs NPH feature importance for V3.

Features with top 30 feature importance values for RF are shown. For each feature, feature importance for both RF and XGBoost are both reported. Features from different visits are encoded in different colors.

**Supplement Figure 5:** PDPs of most significant risk factors identified for sPE vs NPH.

The y-axis is the predictive probability fix at range from 0.46 - 0.56. Confidence intervals are plotted in a dash line. a) PDP for SBP, b) PDP for MAP, and e) PDP for endoglin

**Supplement Figure 6:** PDP of selected top risk factors across 7 different categories that are not presented in Figure 3 or Supplement Figure 6.

The y-axis is the predictive value and the range is different across all plots in order to easily analyze the effect of each feature on predictive probability.
