## Supplementary figures and images for "Preeclampsia Predictor with Machine Learning: A Comprehensive and Bias-Free Machine Learning Pipeline"

### Supplement Figure 1

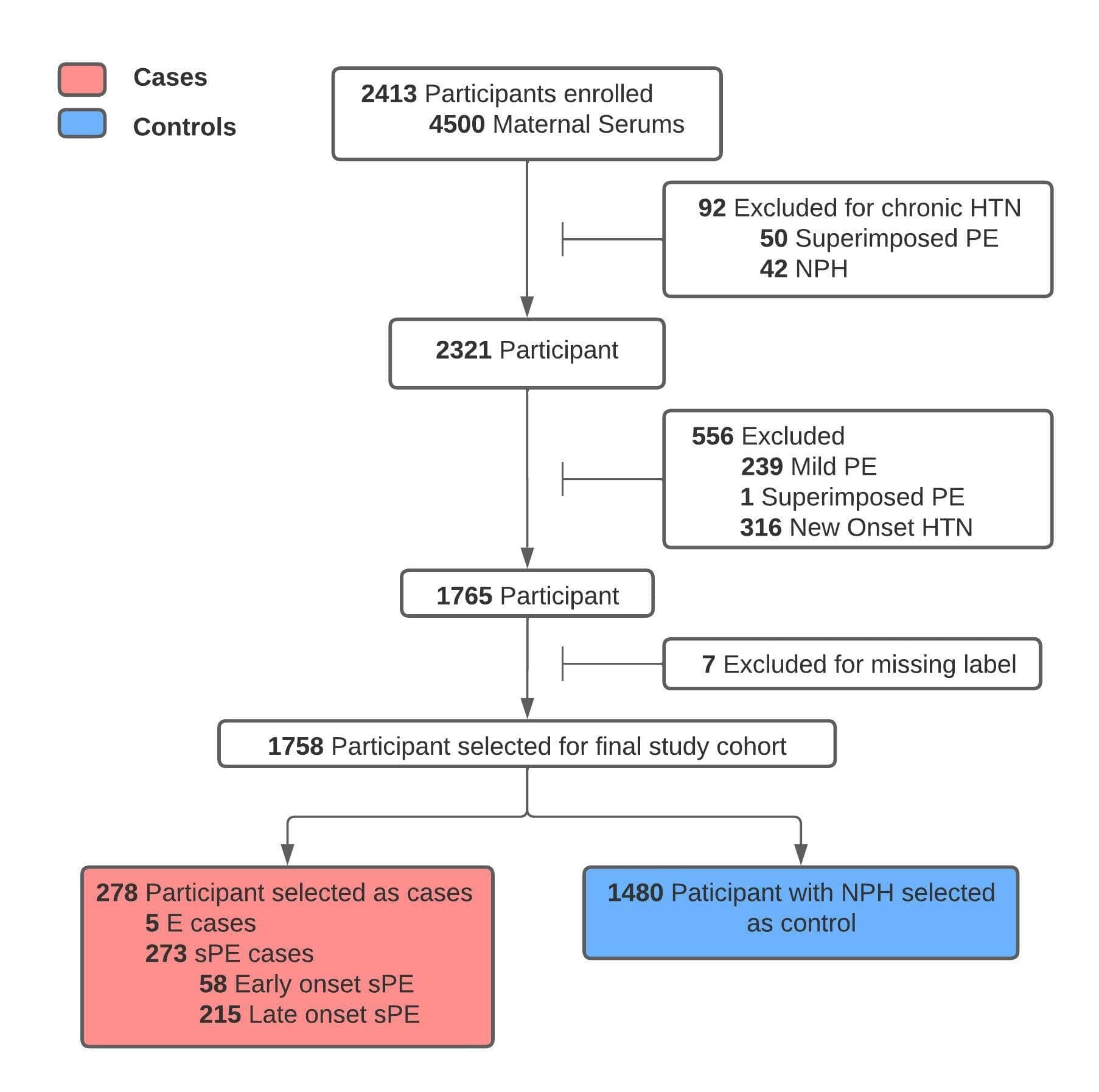

### Supplement Figure 2

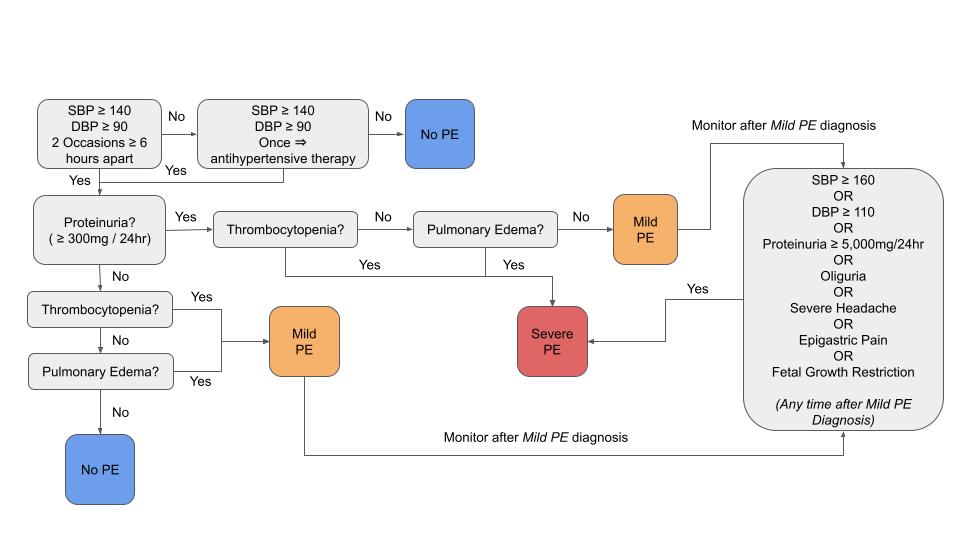

### Supplement Figure 3

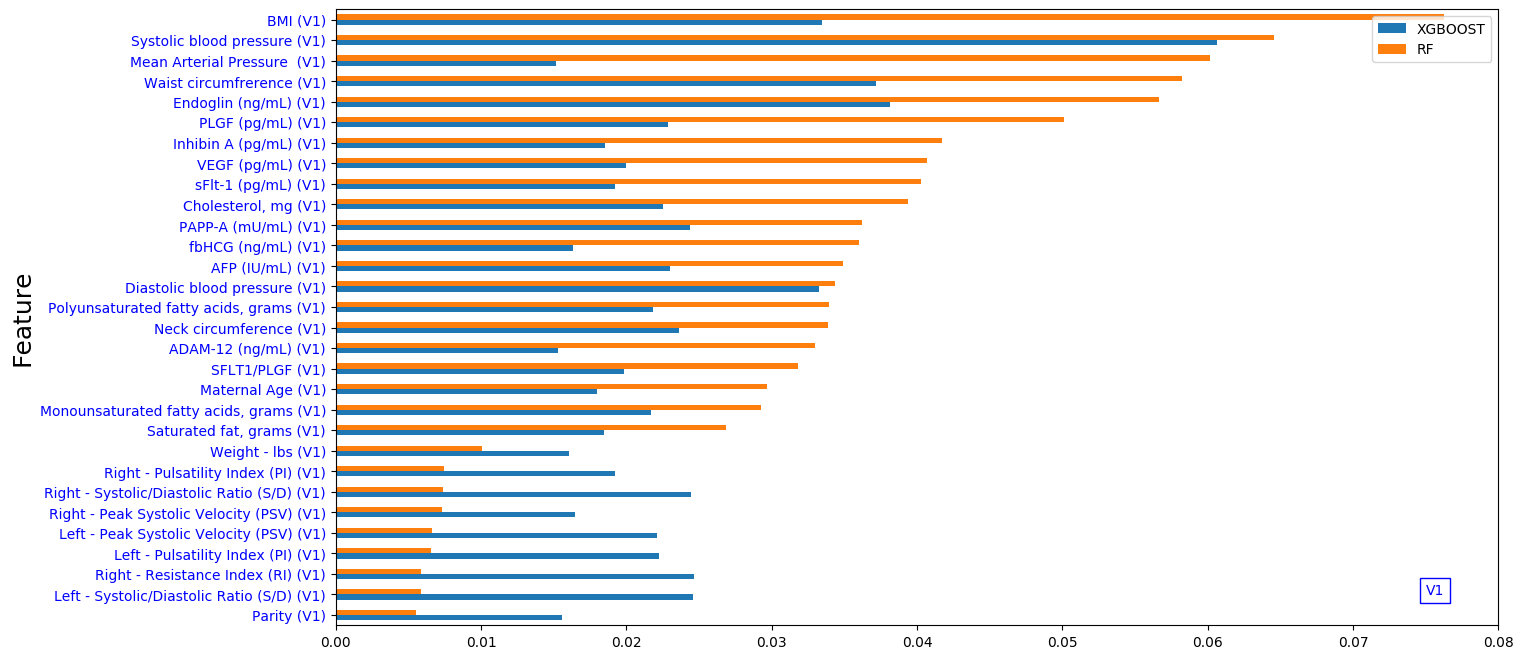

### Supplement Figure 4

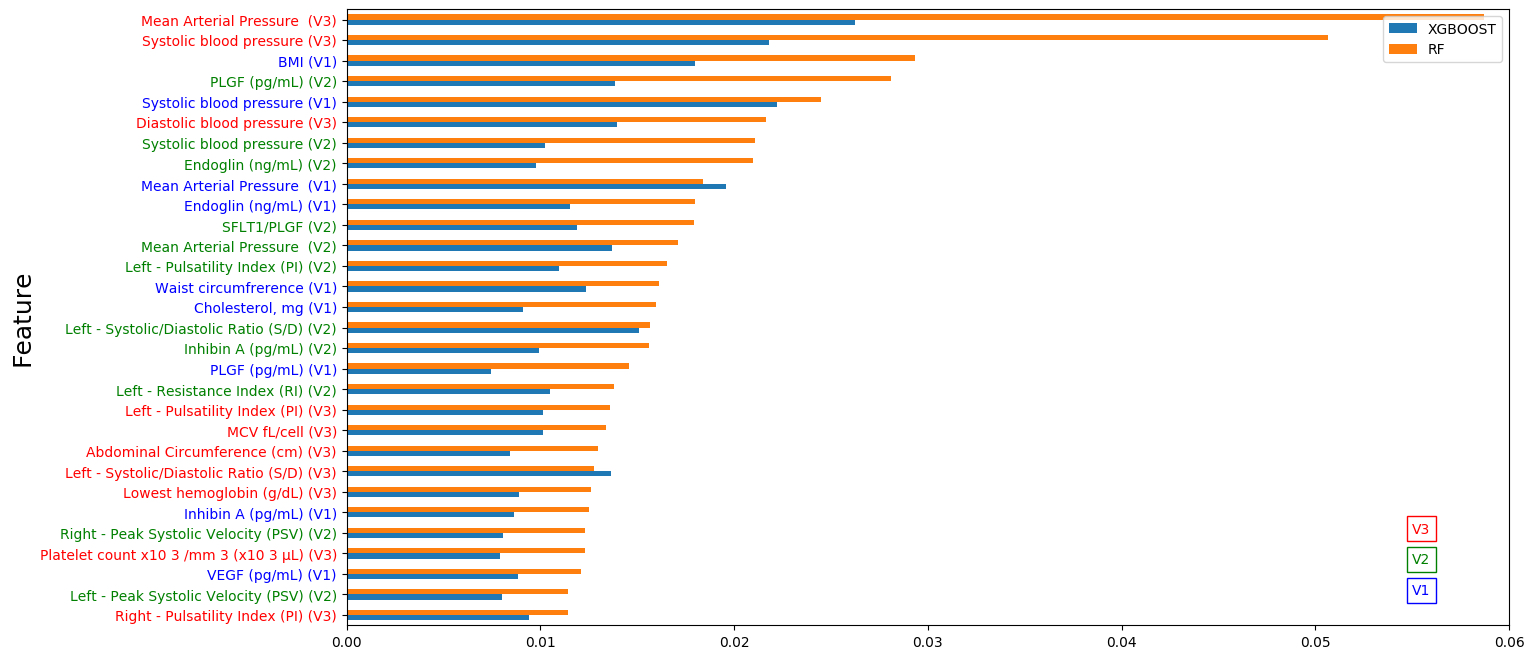

### Supplement Figure 5

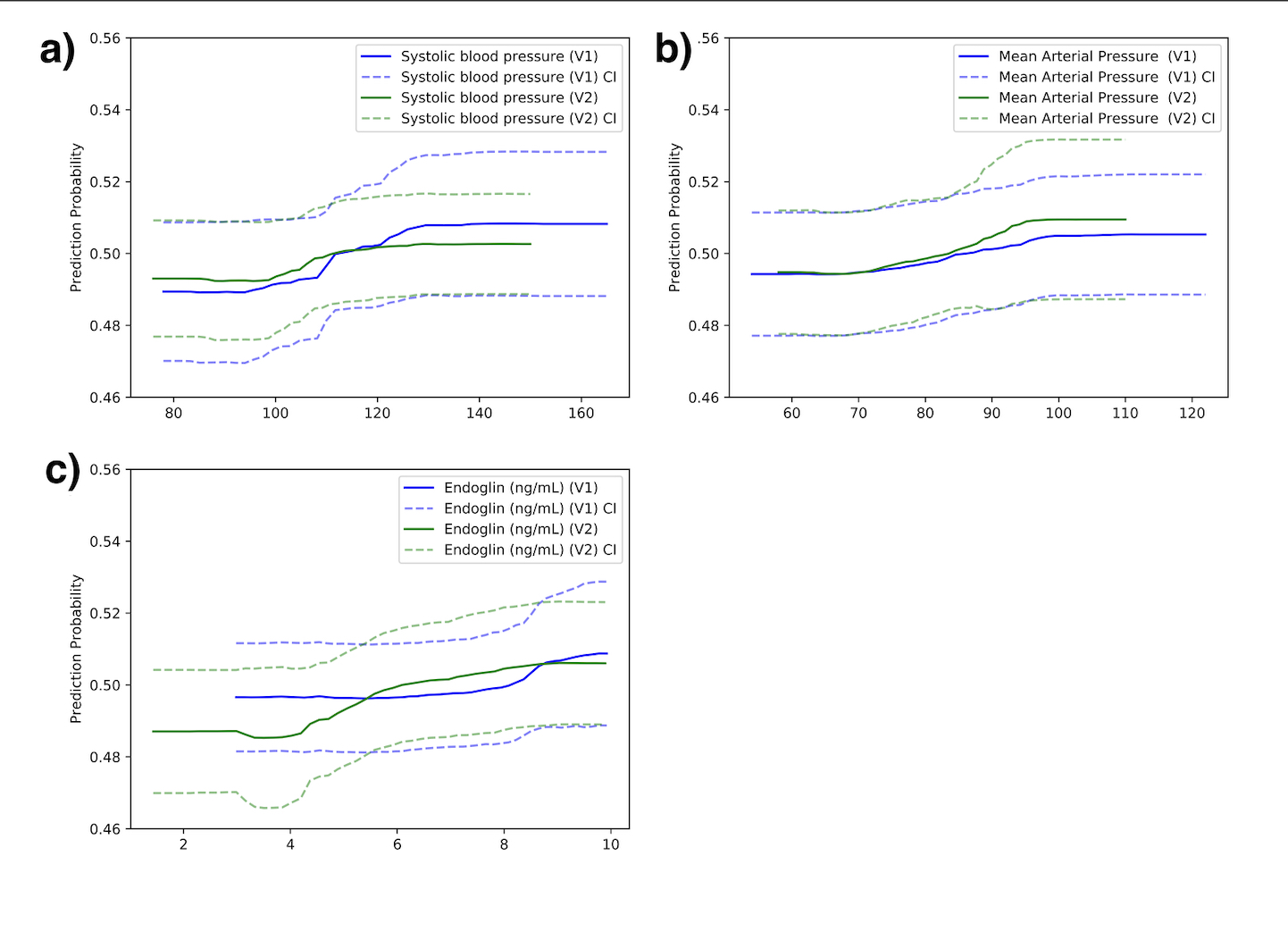

### Supplement Figure 6

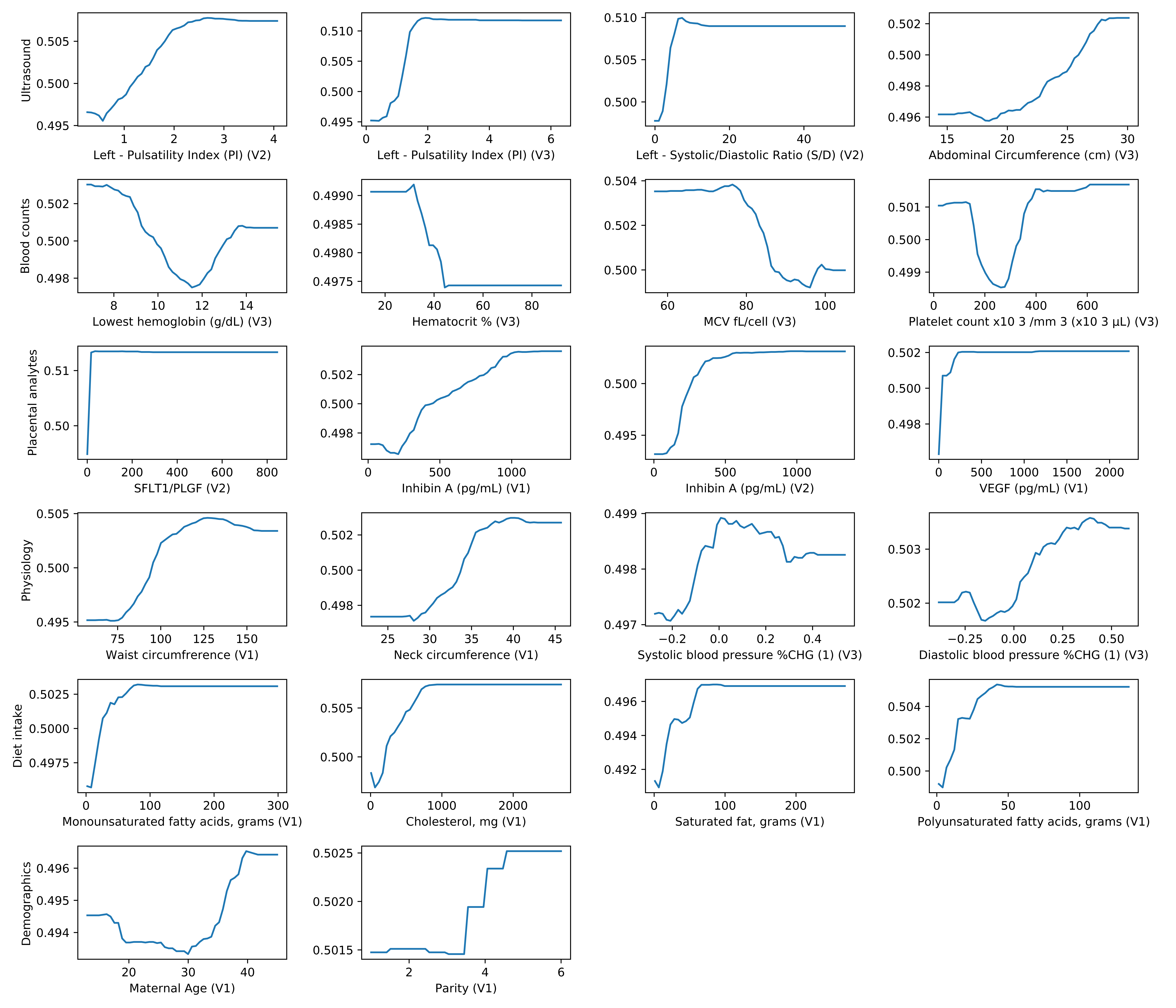
